# A cross-disorder analysis of CNVs finds novel loci and dose-dependent relationships of genes to psychiatric traits

**DOI:** 10.1101/2025.07.11.25331310

**Authors:** Omar Shanta, Marieke Klein, Molly Sacks, Jeffrey R. MacDonald, Adam Maihofer, Mohammad Ahangari, Worrawat Engchuan, Bhooma Thiruvahindrapuram, James Guevara, Oanh Hong, Guillaume Huguet, Ida Sønderby, Maria Kalyuzhny, Mark J. Adams, Rolf Adolfsson, Ingrid Agartz, Allison E. Aiello, Martin Alda, Judith Allardyce, Ananda B. Amstadter, Till F.M. Andlauer, Ole A. Andreassen, María S. Artigas, S. Bryn Austin, Muhammad Ayub, Dewleen G. Baker, Nick Bass, Bernhard T. Baune, Maximilian Bayas, Klaus Berger, Joanna M. Biernacka, Tim Bigdeli, Jonathan I. Bisson, Douglas Blackwood, Marco Boks, David Braff, Elvira Bramon, Gerome Breen, Tanja Brueckl, Richard A. Bryant, Cynthia M. Bulik, Joseph Buxbaum, Murray J. Cairns, Jose M. Caldas-de-Almeida, Megan Campbell, Dominique Campion, Vaughan J. Carr, Enrique Castelao, Boris Chaumette, Sven Cichon, David Cohen, Aiden Corvin, Nicholas Craddock, Jennifer Crosbie, Darrina Czamara, Udo Dannlowski, Franziska Degenhardt, Douglas L. Delahanty, Astrid Dempfle, Guillaume Desachy, Arianna Di Florio, Faith B. Dickerson, Srdjan Djurovic, Katharina Domschke, Lisa Douglas, Ole K. Drange, Laramie E. Duncan, Howard J. Edenberg, Tonu Esko, Steve Faraone, Norah C. Feeny, Andreas J. Forstner, Barbara Franke, Mark Frye, Dong-jing Fu, Janice M. Fullerton, Anna Gareeva, Linda Garvert, Justine M. Gatt, Pablo Gejman, Daniel H. Geschwind, Ina Giegling, Stephen J. Glatt, Joe Glessner, Fernando S. Goes, Katherine Gordon-Smith, Hans Grabe, Melissa J. Green, Michael F. Green, Tiffany Greenwood, Maria Grigoroiu-Serbanescu, Raquel E. Gur, Ruben C. Gur, Jose Guzman-Parra, Jan Haavik, Tim Hahn, Hakon Hakonarson, Joachim Hallmayer, Marian L. Hamshere, Annette M. Hartmann, Arsalan Hassan, Caroline Hayward, Johannes Hebebrand, Sian M.J. Hemmings, Stefan Herms, Marisol Herrera-Rivero, Anke Hinney, Georg Homuth, Andrés Ingason, Lucas T. Ito, Nakao Iwata, Ian Jones, Lisa A. Jones, Lina Jonsson, Erik G. Jönsson, René S. Kahn, Robert Karlsson, Milissa L. Kaufman, John R. Kelsoe, James L. Kennedy, Anthony King, Tilo Kircher, George Kirov, Per Knappskog, James A. Knowles, Nene Kobayashi, Karestan C. Koenen, Bettina Konte, Mayuresh Korgaonkar, Kaarina Kowalec, Marie-Odile Krebs, Mikael Landén, Claudine Laurent-Levinson, Lauren A. Lebois, Doug Levinson, Cathryn Lewis, Qingqin Li, Israel Liberzon, Greg Light, Sandra K. Loo, Yi Lu, Susanne Lucae, Charles Marmar, Nicholas G. Martin, Fermin Mayoral, Andrew M. McIntosh, Katie A. McLaughlin, Samuel A. McLean, Andrew McQuillin, Sarah E. Medland, Andreas Meyer-Lindenberg, Vihra Milanova, Philip B. Mitchell, Esther Molina, Bryan Mowry, Bertram Muller-Myhsok, Niamh Mullins, Robin Murray, Markus M. Nöthen, John I. Nurnberger, Kevin S. O’Connell, Roel A. Ophoff, Holly K. Orcutt, Michael J. Owen, Aarno Palotie, Carlos Pato, Michele Pato, Joanna Pawlak, Triinu Peters, Tracey L. Petryshen, Giorgio Pistis, James B. Potash, John Powell, Martin Preisig, Digby Quested, Josep A. Ramos-Quiroga, Andreas Reif, Kerry J. Ressler, Marta Ribasés, Marcella Rietschel, Victoria B. Risbrough, Margarita Rivera, Alex O. Rothbaum, Barbara O. Rothbaum, Dan Rujescu, Takeo Saito, Alan R. Sanders, Russell J. Schachar, Peter R. Schofield, Eva C. Schulte, Thomas G. Schulze, Laura J. Scott, Soraya Seedat, Christina Sheerin, Jianxin Shi, Pamela Sklar, Susan Smalley, Olav B. Smeland, Jordan W. Smoller, Edmund Sonuga-Barke, David St. Clair, Nils Eiel Steen, Dan Stein, Frederike Stein, Murray B. Stein, Fabian Streit, Neal Swerdlow, Florence Thibaut, Johan H. Thygesen, Ilgiz Timerbulatov, Claudio Toma, Edward Trapido, Micheline Tremblay, Ming T. Tsuang, Monica Uddin, Marquis P. Vawter, John B. Vincent, Henry Völzke, James T. Walters, Cynthia S. Weickert, Lauren A. Weiss, Myrna M. Weissman, Thomas Werge, Stephanie H. Witt, Miguel Xavier, Robert Yolken, Ross M. Young, Tetyana Zayats, Lori A. Zoellner, AGP Consortium, PEIC Psychosis Endophenotypes International Consortium, ADHD Working Group of the Psychiatric Genomics Consortium, Autism Working Group of the Psychiatric Genomics Consortium, Bipolar Disorder Working Group of the Psychiatric Genomics Consortium, Major Depressive Disorder Working Group of the Psychiatric Genomics Consortium, PTSD Working Group of the Psychiatric Genomics Consortium, Schizophrenia Working Group of the Psychiatric Genomics Consortium, CNV Working Group of the Psychiatric Genomics Consortium, Kimberley Kendall, Brien Riley, Naomi R. Wray, Michael C. O’Donovan, Patrick F. Sullivan, Sandra Sanchez-Roige, Caroline M. Nievergelt, Sébastien Jacquemont, Stephen W. Scherer, Jonathan Sebat

## Abstract

Rare copy number variants (CNVs) are a key component of the genetic basis of psychiatric conditions, but have not been well characterized for most. We conducted a genome-wide CNV analysis across six diagnostic categories (N = 574,965): autism (ASD), ADHD, bipolar disorder (BD), major depressive disorder (MDD), PTSD, and schizophrenia (SCZ). We identified 35 genome-wide significant associations at 18 loci, including novel associations in SCZ (*SMYD3, USP7*-*HAPSTR1*) and in the combined cross-disorder analysis (*ASTN2*). Rare CNVs accounted for 1–3% of heritability across diagnoses. In ASD, associations were uniformly positive, consistent with autism having diverse etiologies and clinical presentations. By contrast, CNVs showed a dose-dependent relationship for other diagnoses, including SCZ and PTSD, with reciprocal deletions and duplications having inversely correlated effects and distinct genotype-phenotype relationships. Our findings suggest that genes have effects that are both dose-dependent and pleiotropic, such that a positive influence on one dimension of psychopathology may be accompanied by positive or negative effects on others.

## Introduction

The genetic architecture of neuropsychiatric traits is highly polygenic and consists of a wide range of allelic effects, from common variants of small effect ^1,2^ to rare variants of large effect ^3–5^. In particular, rare copy number variants (CNVs) have provided key insights into the etiology of psychiatric conditions ^3^. Aggregate measures of rare CNV burden provided the earliest evidence for the contribution of rare variants to autism spectrum disorder (ASD) ^6^ and schizophrenia (SCZ) ^7,8^, and studies of rare variants are beginning to make progress in major depressive disorder (MDD) ^9^ and post-traumatic stress disorder (PTSD) ^10^. In addition, genome-wide association studies of rare CNVs have been vital for the discovery of genes and molecular pathways underlying psychiatric conditions. Analysis of CNVs within genes has implicated pathways involved in chromatin regulation ^11^ and synaptic function ^12,13^. Studies in larger samples have found strong associations of specific rare CNVs with ASD ^14,15^ and SCZ ^16–19^. Subsequent whole exome sequencing studies of ASD ^16–18^ and SCZ ^5^ have identified a wider array of rare gene mutations that further implicate pathways in synaptic function and genetic regulation of fetal brain development.

Despite these discoveries, the relationship of genes to the broad spectrum of psychiatric traits is not well defined. No rare variant has yet been found that is specific to a psychiatric diagnosis. Each rare CNV is associated with a variety of mental health and neurodevelopmental conditions ^20^ and cognitive ^21,22^, physical ^23,24^ and general medical ^25^ conditions in the general population. These findings are consistent with significant genetic overlap between diagnostic categories, and are consistent with rare variants being pleiotropic, i.e. each having variable expressivity for multiple psychiatric traits ^26,27^.

Large scale collaborative studies of CNV have the potential to identify novel gene associations and new targets for development of therapeutics. Furthermore, a cross-disorder approach, in which a comparative analysis is done for several psychiatric conditions in parallel, could give a more granular dissection of genotype-phenotype relationships in genes and pathways. Genome-wide association studies (GWAS) are being applied on a large scale to characterize common variant influence and shared genetic risk factors across multiple psychiatric conditions ^28^ in the Psychiatric Genomics Consortium (PGC). Here we apply the large-scale collaborative approach of the PGC to the discovery and characterization of rare variants that influence mental health by genome-wide analysis of CNVs in 574,965 individuals across six diagnostic categories.

## Results

### Rare CNV GWAS identifies 35 genome-wide significant associations at 18 loci

We aggregated microarray intensity files from GWAS datasets to obtain rare CNV calls in 574,965 individuals (**Table S1**) including population controls (N = 441,958) and cases of SCZ (N = 36,865), ASD (N = 13,545), BD (N = 23,119), MDD (N = 38,917), PTSD (N = 17,839), and ADHD (N = 3,544). Individuals spanned multiple ancestries including 513,287 subjects classified as european (EUR), 27,964 asian (ASN/ASAM), 17,606 african (AFR/AFAM) samples, 5,812 Latin-X (LAT), and 10,296 subjects of mixed ancestry (**Table S2**). CNVs were called using a centralized pipeline for systematic CNV calling across genotyping platforms and cohorts. QC of the dataset was performed at the levels of samples (**Fig. S1**), CNV calls, and probes, as described in the methods. Probe normalization methods and CNV calling accuracy are optimal for rare variants and a majority of common CNVs are captured by SNPs ^29^ ; thus, the call set was restricted to CNVs with frequencies <2%.

To identify rare genomic loci that contribute to psychiatric conditions, a CNV GWAS was performed in each group: ASD, ADHD, SCZ, PTSD, MDD, BD as well as in the combined cases, referred to as the cross-disorder group (XD). Statistical association tests were performed for CNV counts in cases and controls at each probe with a CNV across the genome by logistic regression, controlling for cohort, sex and ancestry principal components (PCA in **Fig. S2**). Summary statistics were generated separately for each genotyping platform. Meta-analysis of probe-level summary statistics on CNV associations was performed with METAL ^30^. Parameters for meta-analysis were optimized to control for statistical confounders of CNV GWAS, including heterogeneity of genotyping platforms and sparse data on rare variants (**Fig. S3**). Genome-wide multiple test correction was estimated by permutation in the SCZ cohort to determine the appropriate threshold (Jaccard index) for collapsing correlated adjacent probes to estimate the genome wide correction for each diagnostic category (**Fig. S4**). CNV GWAS was carried out for deletions (DEL) and duplications (DUP) separately in each diagnosis and in the combined XD cohort. Association analyses identified 35 genome-wide significant associations for 18 independent CNV loci (**Fig. 1a, Table S3**).

**Figure 1:**
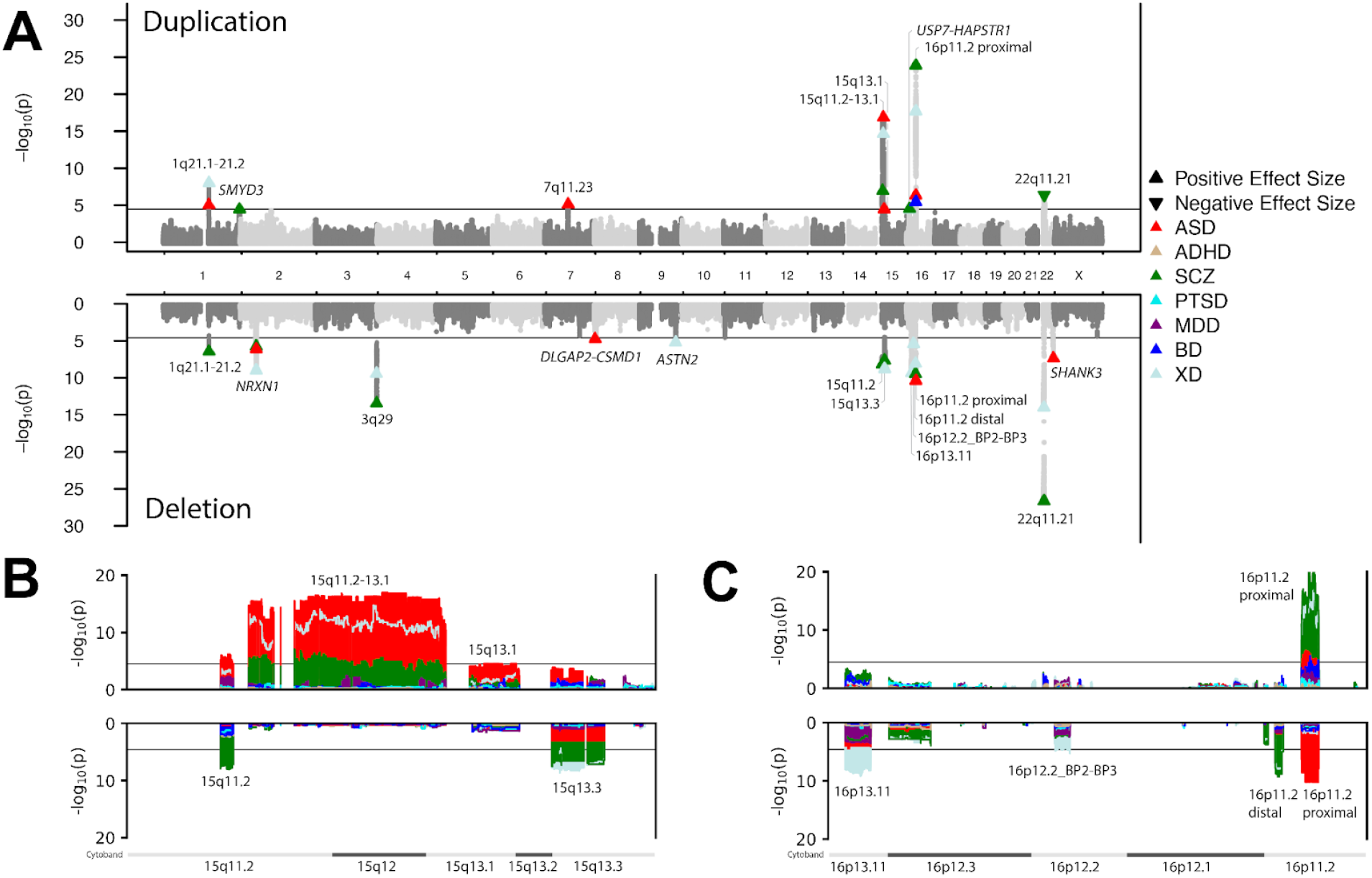
Detection of 35 genome-wide significant associations at 18 loci. A) CNV GWAS was performed at the breakpoint level. CNV GWAS Manhattan plots for all individual disorders were superimposed on top of each other to produce a porcupine plot of all associations in 6 diagnostic categories and in XD. The black line shows the approximate genome-wide significance threshold (DUP: 3.2×10^−5^, DEL: 2.5×10^−5^) since the threshold varies by diagnosis. The genome-wide significance thresholds for the 7 groups are ASD (DUP: 4.9×10^−5^, DEL: 3.5×10^−5^), ADHD (DUP: 4.9×10^−5^, DEL: 3.4×10^−5^), SCZ (DUP: 4.1×10^−5^, DEL: 3.0×10^−5^), PTSD (DUP: 4.7×10^−5^, DEL: 3.4×10^−5^), MDD (DUP: 4.4×10^−5^, DEL: 3.1×10^−5^), BD (DUP: 4.3×10^−5^, DEL: 3.1×10^−5^), and XD (DUP: 3.2×10^−5^, DEL: 2.5×10^−5^). The directionality of effect for each genome-wide significant hit is indicated by upward (positive) and downward (negative) facing triangles. B) Zooming into the cluster of associations on chr15 shows how psychiatric associations differ substantially between CNVs. SCZ is strongly associated with DELs in 15q11.2 and 15q13.3. In contrast, a DUP spanning 15q11.2-13.1 has its strongest association with ASD and to a lesser extent SCZ. C) Zooming into the cluster of associations on chr16 shows how CNV type can result in different psychiatric outcomes within a locus. A DUP at 16p11.2 proximal is strongly associated with SCZ while a DEL in the same location is strongly associated with ASD **Table S3**.

These results demonstrate a broad set of CNV associations that meet genome-wide significance for ASD, SCZ, BD or XD samples. These include many DELs or DUPs that are routinely reported in clinical genetic testing ^31,32^, including CNVs at 1q21.1^33^, 3q29^34^, 16p11.2 ^35^, 22q11.2 ^36^ and several others (**Table S3**). In addition, we find novel associations with CNVs not established previously as causal variants for psychiatric traits. Gene duplications of *SMYD3* were associated with SCZ in this study and have not been described elsewhere in the clinical genetics literature. DELs of *ASTN2* reached genome-wide significance in the XD cases, consistent with *ASTN2* loss of function having a broad association with multiple psychiatric conditions ^37,38^. Duplications of the genes *USP7* and *HAPSTR1* showed a novel association with SCZ in this study.

CNV alleles at five loci showed genome-wide significant associations with more than one diagnosis (1q21.1, *NRXN1*, 15q11.2 BP1-BP2, 15q11.2-13.1, 16p11.2 proximal, **Table S3**) and three reached genome-wide significance only in the XD cohort (*ASTN2*, 16p13.11, 16p12.2 BP2-BP3). These results are consistent with the well-documented pleiotropy of psychiatric risk alleles ^3,20^ and the corresponding overlap in the genetic basis of different psychiatric conditions. ^1,20,28^ For example, a large DUP of 15q11.2-13.1, one of the most well documented rare variants associated with ASD ^39^, is also significantly associated with SCZ (**Fig. 1b, green**), and smaller DELs of this region, 15q11.2 (BP1-BP2/CYFIP1) DEL^19^ and 15q13.3 DEL^8,19^, were also associated with SCZ (**Fig. 1b**). 16p13.11 DEL and 16p12.2 DEL were associated with XD (**Fig. 1c, gray**) while reciprocal DELs and DUPs of 16p11.2 proximal show contrasting associations, the DEL being most significantly associated with ASD ^14^ and the DUP most significantly associated with SCZ ^16^ as well as BD and ASD (**Fig. 1c**).

### Rare CNVs explain 1-3% of the heritability in all diagnostic categories

We and others have shown that the genome-wide burden of CNVs is a significant contributor to ASD and SCZ ^6–8,13^. Conversely, reports of weak associations of CNV burden with BD ^40,41^, MDD^42^, and PTSD ^10^ imply that rare variants could have a comparatively small contribution to the genetic basis of other psychiatric conditions with adult onset. The combined frequency of CNVs at 18 genome-wide significant loci are present in 1.6-3.1% of cases (**Fig. 2A, Table S4**) and 1.4% of controls.

**Figure 2:**
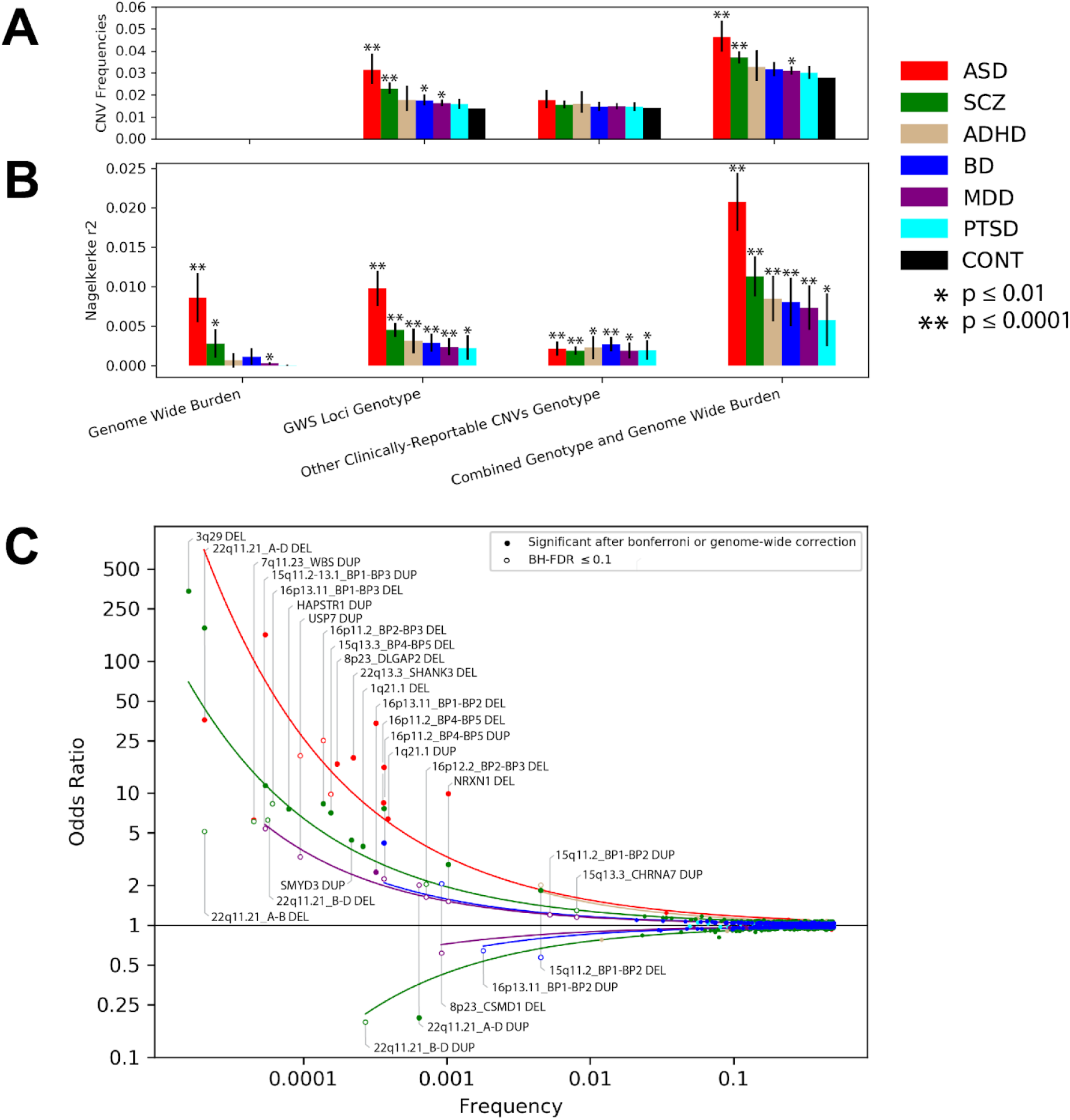
Rare CNVs span a broad range of frequencies and effect sizes, and explain 0.6-2% of the variance in all diagnostic categories. **(A)** Frequencies of CNVs in 6 diagnostic categories, **Table S4**. **(B)** Variance explained by rare CNV genotype (R^2^) in each diagnosis was estimated for the combined loci by logistic regression. Loci were stratified for each diagnosis into genome-wide significant loci, and “other” known pathogenic microdeletion and duplications that are routinely reported in clinical genetic testing but do not reach genome-wide significance in this study. Diagnoses are ordered according to their ranking in the “Combined Genotype and Genome Wide Burden”. Total variance explained by rare CNV was as follows: PTSD (R2=0.58%), MDD (R2=0.74%), BD (R2=0.81%), ADHD (R2=0.85%)m SCZ (R2=1.11%), and ASD (R2=2.08%), results in **Table S5**. **(C)** Effect sizes of significant loci in the CNV-GWAS are shown as a function of frequency. There was an average of 50.7 independent tests in each diagnostic category. Effect sizes are given for all CNVs that show at least one association that meets BH-FDR≤10% correction (open circles) In addition, all associations that meet Bonferroni correction for 50 tests (P≤0.001) or meet genome-wide significance are shown (see Methods) (solid circles). Effect sizes for significant SNPs from previous SNP-GWAS were included to model variation effects across the full frequency spectrum. Results for DEL and DUP effect sizes at 18 loci are in Supplementary **Table S9**.

Estimates of the total CNV burden genome-wide suggest significant disparities between diagnostic categories in the contributions of rare variants. The burden of risk alleles (genome-wide significant) was increased in ASD and to a lesser extent in SCZ, BD and MDD; however the collective frequency of risk alleles in ADHD and PTSD was not significantly greater than in controls (**Fig. 2A**). The total variance explained by CNV burden (length) was estimated by meta-analysis of Nagelkerke’s R^2^ estimates from logistic regression. As expected, variance explained by genome wide burden was greatest for ASD (R^2^=0.86%) and SCZ (R^2^=0.28%) and weaker for BD (R^2^=0.11%), ADHD (R^2^=0.07%), MDD (R^2^=0.03%), PTSD (R^2^=2.78×10^−5^)(**Fig. 2B, Table S5**). When we dissected CNV burden by functional categories of length (bp), large CNVs (>1 Mb), and loss of function intolerance (pLI>0.5), CNV burden for all diagnoses was increased in one or more functional categories (**Fig. S5, Table S6**). We found modest evidence for a reduced CNV burden in loss-of-function intolerant genes in PTSD relative to controls (**Table S6**). This weak effect could imply that some rare variants reduce the probability of a PTSD diagnosis or it might be consistent with case ascertainment of some PTSD cohorts selecting against traits attributable to deleterious variants in genes (such as intellectual disability).

There was less disparity between diagnostic categories in the contribution of rare variants, when we quantified variance explained by CNV genotype rather than their collective burden. Estimation of Nagelkerke’s R^2^ from logistic regression finds that CNVs at genome-wide significant loci explain 0.2-1% of the variance across all 6 diagnostic categories (**Fig. 2B, Table S5**). An additional 0.2-0.3% was explained by other clinically-reportable CNVs ^21,43^ (**Table S7**) that were not genome-wide significant in our study (**Fig. 2B, Table S5**). In total, the variance explained by CNV genotypes and genome-wide burden combined was non-trivial in all disorders and ranged from 0.6% to 2% **(Fig. 2B, Table S5)**. Transformation of the Nagelkerke’s R^2^ to a liability scale gives heritability estimates of 0.9% to 3% (**Table S8**).

When the six diagnoses were ranked based on the variance explained by rare CNVs (ASD > SCZ > ADHD > BD > MDD > PTSD), the R^2^ estimates in this study were correlated with corresponding estimates of heritability from twin studies (Spearman’s rank correlation P = 0.0028, **Table S8**). However, when R^2^ was converted to a liability scale, the correlation with twin heritability was not significant (p = 0.48) and neither estimate was strongly correlated with current estimates of SNP-based heritability (**Table S8**). Given that heritability estimates rely on estimates of diagnosis prevalence that have been changing over time ^44,45^, we regard our estimates of Nagelkerke’s R^2^ to be a more accurate representation of the relative contributions of rare variants across the six diagnostic categories.

### Characterizing pleiotropic effects of CNVs across 6 diagnoses

To gain a more complete view of the spectrum of diagnoses associated with each CNV, we estimated the effect sizes for specific CNV alleles within 18 genome-wide significant loci across 6 diagnostic categories. Within each association peak, distinct CNV alleles with recurrent breakpoints were delineated (**see Table S9**). At association peaks where breakpoints were not recurrent (i.e. were randomly distributed), individual genes were tested (*SMYD3* at 1q44, *ASTN2* at 9q33, *USP7* and *HAPSTR1* at 16p13.2, and *DLGAP2, MYOM2* and *CSMD1* at 8p23). We included all tests for which there were at least 12 CNVs in the combined sample. In total, 50 alleles or genes were tested across 6 diagnoses, and 67 associations were found at a false discovery rate (BH-FDR) ≤ 10%. (**Table S9**). **Figure 2C** displays a trumpet plot of effect size vs frequency, combining rare CNV with common SNP associations from the summary statistics of the PGC GWASs of ASD ^46^, SCZ ^47^, BD ^48^, MDD ^49^, PTSD ^50^, and ADHD ^51^. Curves for each group were fitted to an exponential model to provide a unified representation of rare and common variants. (**Fig. 2C**).

Nearly all loci identified in this study show evidence of association with multiple diagnoses, consistent with genes having pleiotropic effects on psychiatric traits ^20^. ASD is notable for having many rare variants with large effects, all of which are in the positive direction (associated with cases). By contrast, the genetic architecture of other diagnoses consists of a mixture of positive (higher rate in cases) and negative associations (higher rate in controls). The negative associations that were observed, however, do not represent “protection” from all mental health conditions. Without exception, all CNV alleles that had a negative association with one diagnosis also had a positive association with another in this study (15q11.2 BP1-BP2 DEL, 22q11.2 A-D Dup, CSMD1 DEL) or in previous studies (16p13.11 BP1-BP2 DUP^52^, 22q11.2 B-D DUP^53^), consistent with the same gene(s) having divergent effects in different disorders.

### Genes have dose-dependent effects on psychiatric traits

DELs and DUPs of the 22q11.21 A–D region show opposing associations with schizophrenia (SCZ), with DELs increasing susceptibility and DUPs decreasing it ^54^, and both associations reached genome-wide significance in this study. In addition, across all CNVs, the effect sizes of reciprocal DELs and DUPs for SCZ were inversely correlated (**Fig. 3A**), consistent with a linear dose-response relationship. A similar inverse correlation was observed for PTSD (**Fig. S6**). Despite having numerous strong associations, this inverse relationship was not evident for ASD where the DEL-DUP correlation was weakly positive (**Fig. 3B**). The contrasting dose-response curves for SCZ and ASD highlight key differences between these diagnostic categories. With respect to the opposing effects of DEL and DUP, the diagnostic category of SCZ is generally associated with one or the other, whereas the diagnostic category of ASD is often associated with both.

**Figure 3:**
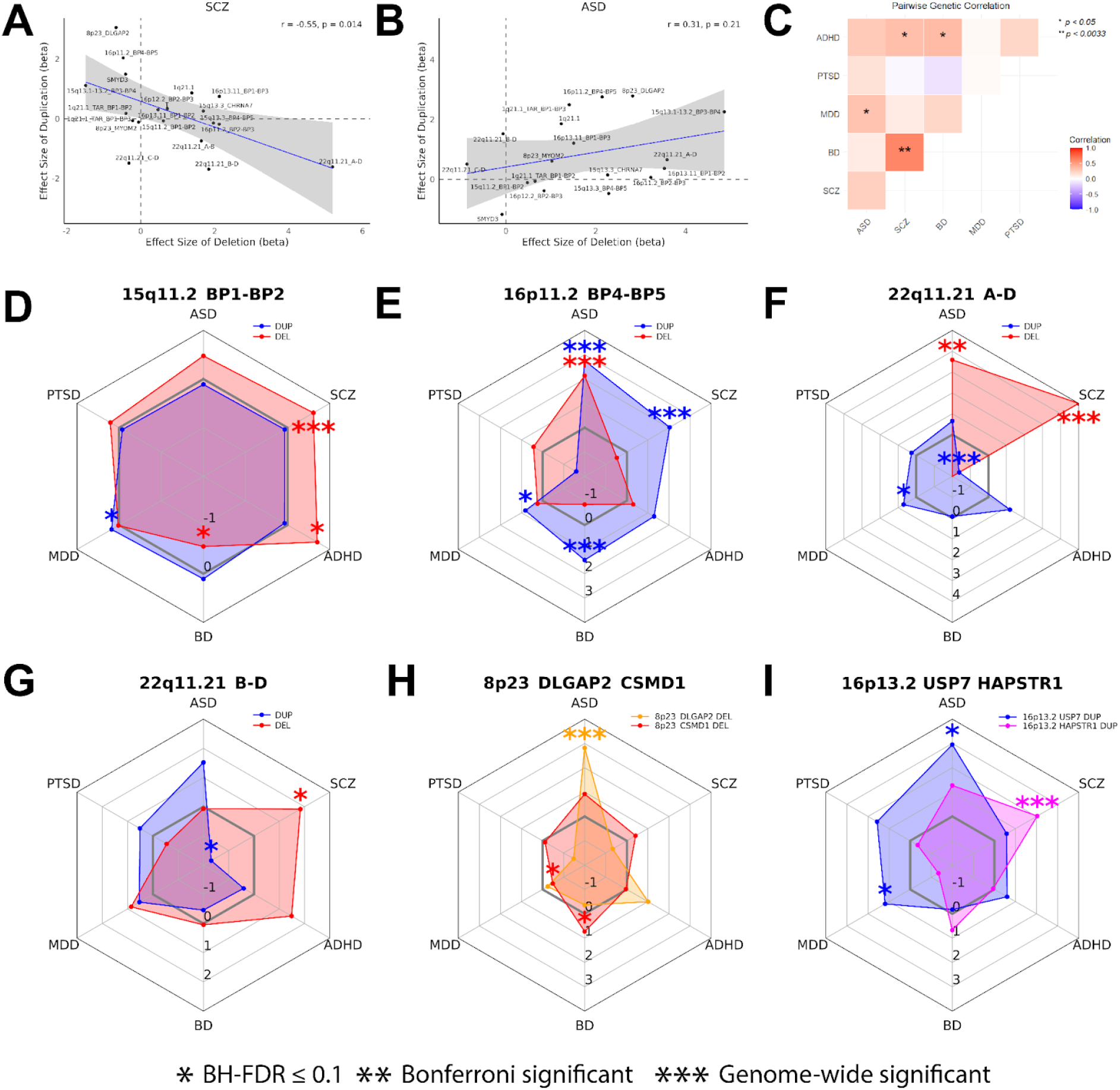
Dose response curves show inverse correlation of effect sizes for deletion and duplication in (A) SCZ, but not (B) ASD. (C) Pairwise genetic correlations of six disorders based on CNV associations, see **Table S10**. (D-G) Effect sizes of deletion and duplication are displayed for each diagnosis or (H-I) Effect sizes for 2 different genes across diagnoses under a single association peak. *BH-FDR<10%; **Bonferroni correction for 50 tests (p<0.001); ***Genome-wide significance. Radar plots for all loci are shown in **Figure. S7** and summary statistics of effect sizes are in **Table S9**.

Analysis of the pairwise genetic correlations of diagnostic categories based on CNV effect sizes (**Table S9**) shows a significant genetic correlation of SCZ and BD (**Fig. 3C, Table S10**, p=3.32×10^−6^). Weak correlations were also observed between these disorders and ADHD (BD p-value=0.01, SCZ p-value=0.02) and between MDD and ASD (p=0.03). The genetic correlations that we observe for CNVs are consistent with genetic correlations observed in GWAS ^1,28^. The notable absence of correlation for SCZ and PTSD highlights how the dose-dependent relationships of genes is evident for both disorders (**Fig. S6**) despite having different associations with specific loci.

While nearly all CNVs were associated with multiple diagnoses, this was not a reflection of rare variant effects being highly non-specific. To the contrary, different alleles exhibited different spectra of associations. Some loci show contrasting phenotype associations for reciprocal DEL and DUP of the same genes including 15q11.2 BP1-BP2 (**Fig. 3D**), 16p11.2 BP4-BP5 (“proximal 16p”, **Fig. 3E**), 22q11.2 A-D (**Fig. 3F**) and 22q11.2 B-D (**Fig. 3G**). Results are consistent with these CNVs having dose-dependent effects on psychiatric traits, and within the 22q11.21 CNV, the dose-dependent effect may be driven by genes within the B-D locus.

At other loci where breakpoints were not recurrent, we compared the effect sizes for multiple genes within the same association peak. Within 8p23 for example, DELs of *DLGAP2* have a strong association with ASD, while DELs of nearby *CSMD1*, a gene that has been previously implicated in GWAS ^2,55^ show contrasting associations with mood disorders BD and MDD (**Fig. 3H**). Within 16p13.2, DUPs of *USP7* are associated with ASD and MDD, while the genome-wide significant association of DUPs with SCZ appears to be strongest for the adjacent *HAPSTR1* gene (**Fig. 3I**). Radar plots of effect sizes for DELs and DUPs across 6 diagnostic categories are shown for all loci tested (**Fig. S7, Table S9**).

Relationships of CNV genotype to phenotype that are observed here are attributable to the dose-dependent effects of genes on pathways and cellular processes As described in our companion paper ^56^, the diagnostic categories of schizophrenia, autism and mood disorders can be differentiated based on the gene-dosage effects on pathways stratified by cell-type and brain region. In particular, 16p11.2 BP4-BP5 and 22q11.2 A-D are enriched for distinct cellular processes that are characteristic of the DUP and DEL effects respectively that contribute to SCZ. Thus the spectrum of clinical phenotypes observed for CNVs in Figure 3E-F is a reflection of how pathway effects are concentrated in neural cell types and cortical brain regions ^56^.

### Recurrent mutations in large neural genes highlight novel associations

Without exception, all associations found in this study occurred in genomic regions that are prone to high rates of structural mutation. Twelve loci consist of hot-spots for non-allelic homologous recombination (NAHR)^57^ that have recurrent breakpoints, in most cases span multiple genes, and are given a cytoband-breakpoint label in **Figure 2** (e.g. 1q21.1) to reflect the CNV allele that was tested. The remaining six loci were common fragile sites (CFS) where double strand breaks occur with high frequency and are distributed more randomly. ^58^ CFS loci are labeled with gene symbols to reflect the genes that were tested (*ASTN2, NRXN1, SMYD3, SHANK3, DLGAP2-CSMD1, USP7-HAPSTR1*, **Table S3**). The fact that all associations occur within CNV hotspots is consistent with the statistical power for rare variants being greatest for the loci with the highest mutation rates ^59^.

All three single-gene associations consist of fragile sites within long genes that have functions related to neuronal development ^58^. Long genes are prone to replication stress, and consequently, genome instability ^60^, and tend to co-localize with TAD boundaries^61^. These include positive associations of *NRXN1* DELs in SCZ (**Fig 4A**), *ASTN2* DELs in XD (**Fig. 4B**) and *SMYD3* DUPs in SCZ (**Fig 4C**). CNVs in these genes have a characteristic fragile-site signature where the breakpoints, lengths and functional consequences of the CNVs are variable. When we stratified CNVs by predicted functional consequence, the associations of *NRXN1* and *ASTN2* DELs were greatest for loss-of-function (LoF) variants that are predicted to result in truncation of the protein (**Table S11**). The association of *SMYD3* DUPs was driven by variants that span at least one full length transcript of the gene, consistent with a gain of function effect (**Table S11**).

**Figure 4:**
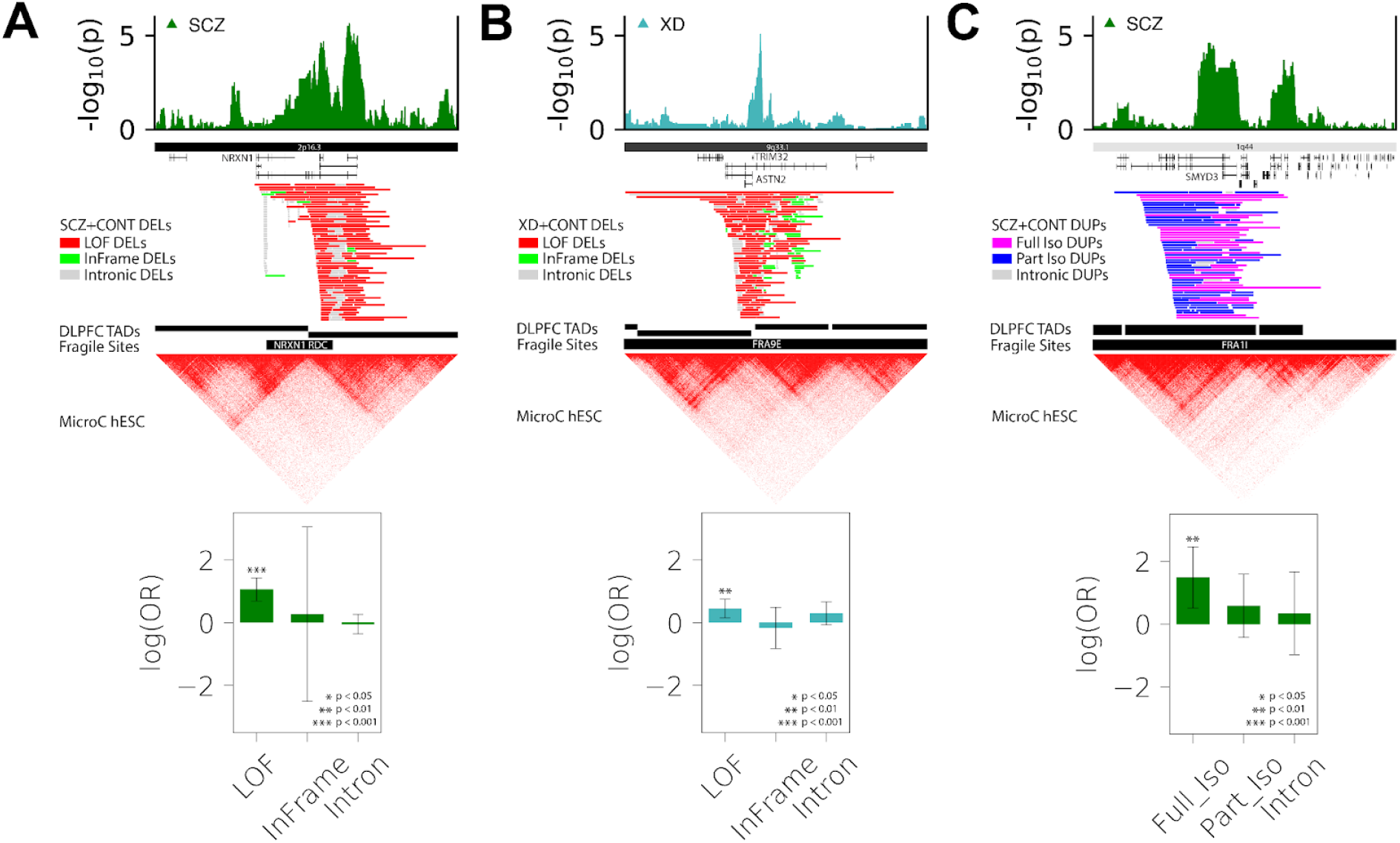
CNV associations implicate large neural genes within common fragile sites. Three genome-wide significant loci that overlap fragile sites and TAD boundaries from Dorsolateral Prefrontal Cortex (DLPFC TADs) are shown for A) NRXN1 B) ASTN2 and C) SMYD3. Association tests (bottom) were stratified by predicted functional consequence: loss of function (LOF), In frame, and intronic (Intron). Duplications spanning a full length isoform (Full Iso) or partially spanning an isoform (Part Iso), and intronic (Intron). **Table S11**.

### Recurrent duplications of *SMYD3* are associated with schizophrenia

The association of the gene SET and MYND domain containing 3 (*SMYD3*) with SCZ appears to be driven by DUPs of full length transcripts, suggesting that the causal variants increase the number of functional copies of *SMYD3*. However, gene duplications detected by microarray can have hidden complexity that, in some cases, results in loss rather than gain of function^62^. To clarify the structure and functional consequence of *SMYD3* DUPs, DNA samples from 3 carriers with DUPs spanning the short isoform of *SMYD3* were obtained from DNA samples available to our group through UCSD and collaborators at Trinity College Dublin, and HiFi long-read whole genome sequencing was performed on each sample using the Pacific Biosciences Revio platform to a total coverage of >20X (**Fig. 5A, Table S12 for coverage and QC**). HiFi long reads were aligned to both the GRCH38 and CHM13 assemblies using PBMM2, and SVs were called using Sniffles and LUMPY. In each genome, DUP breakpoints were identified and contigs were assembled from breakpoint-spanning reads using Flye v2.9.2. Assembled breakpoints revealed that each SV had a distinct structure. Subject 1 carried a tandem duplication of 666,289 bp spanning the short isoform of *SMYD3* (ENST00000403792.7, **Fig 5B**). Subject 2 carried a 516,050 bp non-tandem duplication of ENST00000403792.7 that was inserted into the first intron of *OR2G6* (**Fig 5C**). Subject 3 carried a tandem duplication of 1.6 Mb spanning >10 genes including the full length of the *SMYD3* gene (**Fig 5D**). Thus, all DUPs appear to result in an increased copy number of at least one full length isoform of *SMYD3*.

**Figure 5:**
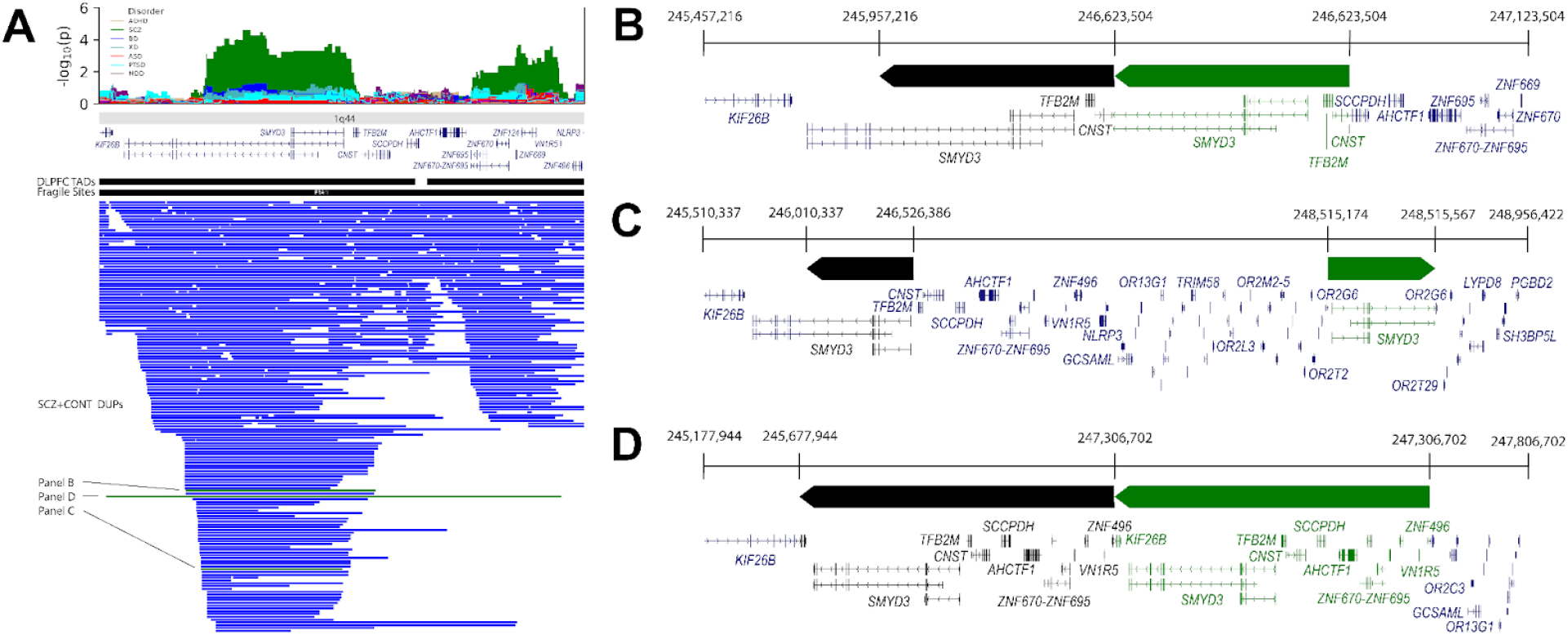
Structures of SMYD3 duplications resolved by HiFi long read WGS. (A) The association peak spans exons 1-3 of the SMYD3 gene while the CNVs themselves have variable breakpoints that duplicate different nearby genes. Pacbio sequencing was performed on 3 cases with the SMYD3 Dup and the corresponding microarray CNV calls are shown (green) on the SCZ DUPs track. (B) The first sample is a tandem duplication that spans the full short isoform of SMYD3. The duplicated portion truncates the long isoform of SMYD3 on the proximal side as well as the CNST gene on the distal side. The inserted sequence has portions of both CNST and SMYD3 sitting flush with each other. (C) The next sample is an inserted DUP that spans the full short isoform of SMYD3. It does not include any extra genes and is inserted next to the olfactory receptor genes with a small deletion of 393bp. (D) The last sample is another tandem duplication that spans all isoforms of SMYD3. It is a longer DUP that includes many genes on the distal side of SMYD3. This DUP cuts KIF26B on the proximal side of SMYD3 and ZNF496 on the distal side and the resulting genome in the sample has both genes sitting flush with each other. **Table S12**.

All breakpoint positions were non-recurrent (unique to each subject). In two cases, tandem duplications resulted in the partial duplication and recombination of genes near the breakpoint junction. However, the fused transcript pairs *CNST*/*SMYD3* (**Fig 5B**) and *KIF26B*/*ZNF496* (**Fig. 5D**) were on opposite strands. In patient 3, the *SMYD3* gene is inserted within *OR2G6* (**Fig 5C**) in the same orientation, and is predicted to fuse exons 1-5 of the long isoform of *SMYD3* with the full open reading frame of *OR2G6*. Thus, in addition to duplicating the short isoform of *SMYD3*, the SV has the potential to cause ectopic expression of an olfactory receptor from the *SMYD3* promoter.

## Discussion

In a comparative study of rare CNVs across six psychiatric diagnoses (ASD, ADHD, SCZ, BD, MDD, and PTSD), rare variant associations elucidate key aspects of the genetic architecture. This study identified 35 genome-wide significant associations at 18 different CNV regions, providing a map of loci where gain or loss of gene dosage influences psychiatric traits. This represents a 4-fold increase in the number of CNV associations to date that meet genome-wide significance in the PGC ^13^.

Rare CNVs accounted for 0.6–2% of the variance in case status across 6 diagnostic categories, corresponding to 0.9–3% of liability-scale heritability. This contribution is independent of the heritability explained by common variants. ASD is notable for having many rare variants with large effects, all of which are in the positive direction. For other diagnostic categories, such as SCZ, BD and MDD (Fig. 2C), the contribution of rare variants consisted of a mixture of risk alleles (more frequent in cases) and protective alleles (more frequent in controls). Hence, the genome-wide burden of rare variants in a categorical diagnosis is not a particularly accurate measure of their influence. The collective frequency of rare CNVs in some disorders, such as ADHD and PTSD, is similar to the frequency in controls but CNVs still account for approximately 1% of heritability.

Importantly, the negative associations that we observe do not represent broad “protection” from mental health conditions. Without exception, all CNV alleles that had an effect size that was negative for one diagnosis have a positive association with another in this study (15q11.2 BP1-BP2 DEL, 22q11.2 A-D DUP, CSMD1 DEL) or in previous studies (16p13.11BP1-BP2 DUP^52^, 22q11.2 B-D DUP^53^), highlighting how the same CNV allele can have divergent, sometimes opposing, associations with different psychiatric traits.

Furthermore, different CNV alleles have different spectra of associations across 6 diagnoses. These contrasts are particularly evident for reciprocal DEL and DUP of the same genes (**Fig. 3**). Previous studies have shown that CNVs have “mirror” dose-dependent effects on a variety of complex traits ^23,24^, including brain structure ^63^ and cognition ^64,65^. Here we show that this principle also applies to psychosis and other psychiatric traits. Reciprocal DELs and DUPs of 22q11.21 show opposing positive and negative associations with schizophrenia (SCZ) respectively ^54^. Furthermore, across all loci, there was an inverse correlation of effect sizes for reciprocal CNVs in SCZ and PTSD, suggesting that CNVs have a dose-dependent relationship with some diagnoses. ASD, by contrast, was characterized by a neutral (weakly positive) dose-response curve. Thus, with respect to the opposing effects of DEL and DUP, SCZ is generally associated with only one of the two extremes, but both often fall under the diagnostic umbrella of ASD. MDD showed a similarly neutral dose-response curve (**Fig. S6**), and ASD and MDD exhibited significant genetic correlation (**Fig. 3C**). These findings underscore how mental health traits related to social behavior and mood are characterized by substantial clinical and genetic heterogeneity ^66^. Deeper clinical characterization of dimensional cognitive phenotypes associated with reciprocal CNVs could better elucidate relationships of gene dosage with quantitative traits and could help to dissect the clinical and genetic heterogeneity within these diagnostic categories ^67^.

Multiple novel loci were identified in SCZ (*SMYD3, USP7*-*HAPSTR1*) and in the combined XD sample (*ASTN2*). SET and MYND domain-containing protein 3 (*SMYD3*) is a histone methyltransferase expressed predominantly in the brain ^68^ with a function in chromatin regulation ^69^. While it’s role in regulation of neural function is not known, a recent study has shown that *SMYD3* expression is elevated in the prefrontal cortex of patients with Alzheimer’s disease, and a SMYD3 inhibitor rescues synaptic and cognitive deficits in a mouse model of tauopathies ^70^. A variety of inhibitors of *SMYD3* have been developed in oncology ^71^. Thus, validation of increased *SMYD3* dosage as a risk factor for psychosis could offer a potential avenue for development of new therapeutics. DUPs that span *USP7* and *HAPSTR1* were associated with SCZ in this study. More detailed characterization found evidence that the *USP7* gene was associated with ASD and MDD, consistent with recurrent *de novo* DUPs of this region previously reported in ASD ^72^. Ubiquitin-specific protease 7 (*USP7*) is a deubiquitinating enzyme that is highly expressed in neuronal cells in cerebrum, cerebellum, and hippocampus^73^. *USP7* influences neuronal development and function through neuronal migration ^74^, dendritic spine morphogenesis^75^ and neuroinflammation.^76^ The SCZ association with this locus was strongest for the adjacent gene *HAPSTR1* (**Table S9**), which encodes a regulator of stress response pathways ^77^, which have been implicated in the etiology of SCZ ^78^. DELs of *ASTN2* were significant in the combined XD cases suggesting these confer risk for a broad range of psychiatric disorders. This finding is consistent with previous reports of *ASTN2* DELs in small samples of SCZ, BD and ASD ^37,79^. *ASTN2* is predominantly expressed in the brain and plays a role in neuronal migration ^80^ and synaptic function ^81^. Common SNPs within *CSMD1*^*82*^ and *NRXN1*^*2*^ are associated with SCZ, and *ASTN2* has been implicated in GWAS of MDD^83^ and BD^84^. Thus, a convergence of evidence from both rare and common variants implicate the same genes. Synaptic neurotransmission, regulation of synaptic plasticity, chromatin and post-transcriptional regulation are common threads between studies of CNVs, whole exome sequencing ^5^ and GWAS ^85^. A more detailed characterization of the molecular pathways, cell types and brain regions that are implicated by CNV associations is described in our companion paper ^56^.

A majority of the CNVs that reached genome-wide significance in this study are variants that are routinely reported in clinical microarray (CMA) testing, and clinically-reportable CNVs that did not reach genome-wide significance in this study explained another 0.2-0.3% of the variance across all six diagnoses (see **Table S5**). Yet very few of the subjects enrolled in this study have been offered genetic testing as part of their clinical care. CMA has historically been restricted to pediatric populations for the evaluation of congenital malformations^86^, intellectual disability and autism ^87^, and insurance providers approve it only for these indications. Consequently, the clinical features of CNVs that are described in the literature reflect this bias, and may not be representative of clinical presentations of CNV carriers in the broader adult population ^25,88^. As large-scale studies like this one begin to uncover associations between CNVs and adult-onset health conditions, the effectiveness of early tailored interventions can be evaluated, clinical guidelines can be revised, and the need for genetic counseling in subjects carrying these CNVs can be assessed. When considering all clinically-reportable variants that influence mental health, the overall difference in diagnostic yield between different clinical populations is relatively small (**Fig. 2A**). If the rationale for ordering a clinical genetic test is the utility of genetic information for lifelong clinical management and genetic counseling of individuals, then the utility of CMA is not limited to the pediatric developmental clinic.

## Data Availability

All data produced in the present study are available upon reasonable request to the authors

## Acknowledgements

We thank the many research participants. The PGC is supported by grants to UCSD (R01MH124847), UNC (R01MH124871), MGH (R01MH124851), Mount Sinai School of Medicine (R01MH124839), Cardiff University (R01MH124873), Trinity College Dublin (R01MH124875) and Washington University St. Louis (R01DA054869). Additional support was provided by grants to J.S. (MH119746, MH133899), C.N. (MH106595, MH124847) and S.J. (U01 MH119690, U01 MH119739, CIHR_495906). Funding for the work in Bipolar Disorder was supported by the Research Council of Norway (#223273, 248778, 262656, 273291, 283798, 248828), South East Norway Health Authority (2017-112), and KG Jebsen Stiftelsen. The iPSYCH project is supported by grants from the Lundbeck Foundation (R165-2013-15320, R102-A9118, R155-2014-1724 and R248-2017-2003) and the universities and university hospitals of Aarhus and Copenhagen. Genotyping of iPSYCH samples was supported by grants from the Lundbeck Foundation, the Stanley Foundation, the Simons Foundation (SFARI 311789 to M.J.D.), and NIMH (5U01MH094432-02 to M.J.D.). The Danish National Biobank resource was supported by the Novo Nordisk Foundation. Data handling and analysis on the GenomeDK HPC facility was supported by NIMH (1U01MH109514-01 to A.D.B.). High-performance computer capacity for handling and statistical analysis of iPSYCH data on the GenomeDK HPC facility was provided by the Center for Genomics and Personalized Medicine and the Centre for Integrative Sequencing, iSEQ, Aarhus University, Denmark (grant to ADB).

## Data and code Availability

A WDL workflow containing all steps of CNV calling, QC and CNV-GWAS and meta-analysis code is under construction and will be released on the PGC CNV Github in conjunction with this publication (https://github.com/orgs/psychiatric-genomics-consortium/teams/cnv).

Meta-analysis of summary statistics for 6 diagnoses and the combined XD sample will be released through the PGC downloads page https://pgc.unc.edu/for-researchers/download-results/

Raw genotype and intensity files are available on subset of the cohort PGC dbGAP datasets https://www.ncbi.nlm.nih.gov/projects/gap/cgi-bin/collection.cgi?study_id=phs001254.v1.p1

Simons Foundation Autism Research Initiative SFARI (SSC and SPARK) https://base.sfari.org/

## Supplementary Materials and Methods

## Methods

### GWAS datasets of the PGC

The CNV subgroup of the Psychiatric Genomics Consortium (PGC) works in collaboration with principal investigators from many institutions to obtain large sample sizes of microarray data and analyze them using a centralized pipeline. We acquired microarray intensity files from GWAS for a total of 574,965 samples that included data from 6 psychiatric conditions (**Table S1**). These samples were genotyped on 25 platforms across 4 genome builds. Data from Illumina was collected as either raw intensity data (IDAT) files or final report files while data from Affymetrix was collected as CEL files. To harmonize data, probes for newly acquired datasets were lifted over to GRCH38 for CNV calling while previously called CNVs were lifted over to GRCH38. Samples were genotyped on either Illumina or Affymetrix array.

### Copy number variant calling

For samples that were provided as IDAT files, the Illumina command line version of Genome Studio was used in conjunction with platform-specific manifest and cluster files to produce genotype call (GTC) files. Relevant features were extracted from GTC files to obtain final report files with probes, genotypes, Log R Ratio (LRR), and B Allele Frequency (BAF) for each sample. For samples that were not mapped to GRCH38, probe genome positions were converted to hg38 using the LiftOver tool. Samples within each platform were grouped into batches by plate. For Affymetrix 6.0 arrays, CNVs were called using four methods: PennCNV, iPattern, CScore, and Birdsuite. For Affymetrix 5.0 and 500K arrays, CNVs were called using two methods that were compatible with this platform: PennCNV and Birdsuite. For Axiom arrays, CNVs were called using PennCNV and QuantiSNP. For all Illumina arrays, CNVs were called using two methods: PennCNV and iPattern. The consensus of CNV calls from multiple callers was created by merging CNVs at the sample level and retaining CNVs that were called by at least 2 methods.

### Quality control of samples and CNVs

#### Sample QC

Quality control (QC) was performed first at the sample level according to methods from our previous CNV GWAS of schizophrenia ^13^. For Illumina arrays, LRR standard deviation, BAF standard deviation, and GC waviness factor were extracted from PennCNV log files (**Fig. S1**). Samples were retained if each of the measures were within 3 SD of the median. Affymetrix arrays used MAPD and waviness-sd parameters from affy power tools. The proportion of the chromosome that was tagged as a CNV was calculated and samples were excluded if >10% of the chromosome was marked as a CNV region to filter possible aneuploidies. Distribution of QC metrics differ by genotyping platform (**Fig. S1**), which provides a rationale for performing meta-analysis of summary statistics by platform.

#### CNV QC

A basic set of QC filters were applied to the call set, and subsequent filtering was performed at the probe level. CNVs that were called with different CNV types from different callers were excluded. Large CNVs that were fragmented were merged if one of the calling methods detected a CNV spanning the gap. CNVs < 10kb in length or contained < 10 probes were excluded. CNV calls were removed if they spanned the centromere or telomere (100kb from end of chromosome) or had >50% overlap with segmental duplications, immunoglobulin, or T cell receptor. Because the normalization of microarray intensity data is performed within a batch (typically a 96-well plate), CNV calling is optimal for rare CNVs that show distinguishable deviation in probe intensities relative to other samples within a plate. For common copy number polymorphisms (frequencies >10%), there is high variance in probe intensities within plates as well as sampling variance in the distribution of copy numbers. For this reason, CNV calling accuracy is suboptimal for common CNVs. Furthermore, a majority of common CNVs are tagged by adjacent SNPs ^29^ and are already captured by GWAS. Thus our final call set was restricted to CNVs with ≤ 10% frequency within-platform or across all platforms.

### Probe QC

Our CNV-level QC applied a liberal threshold of 10% frequency to make certain that pathogenic CNVs near the rare CNV frequency boundary were kept in our analysis. CNV frequency was then calculated for each probe and probes with rare CNVs were kept if they contained ≤ 2% CNV frequency within-platform and within-dataset. Additionally, probes were removed if they were heavily filtered by CNV QC (filtered CNVs > 20). Platform-specific and dataset-specific CNVs arose from the differing probe content of the many genotyping arrays. We applied probe-level specificity filters to prevent these CNVs from causing spurious associations in our results as described in detail in the Methods.

### Ancestry principal components and ancestry partitioning

We extracted a subset of SNPs with < 1% missingness across all platforms (12,185 SNPs) and performed a principal component analysis using the flashPCA software ^89^. In order to genetically infer the ancestry of each individual, we used the SNPweights software ^90^ on the same subset of SNPs to calculate % ancestry based on a reference panel containing 6 different populations (751 EUR, 687 EAS, 630 SAS, 568 AFR, 41 AMR, 22 OCE). Samples were categorized into 5 large homogeneous groupings based on the following criteria used in a previous study ^50^ (**Table S2**, **Fig. S2**): EUR: subjects with EUR ≥ 90%, AFR/AFAM: subjects with EUR < 90% & AFR ≥ 5% & EAS/SAS/AMR/OCE < 5%, ASN/ASAM: subjects with EUR < 90% & (EAS ≥ 5% or SAS ≥ 5%) & AFR/AMR/OCE < 5%, LAT: subjects with EUR < 90% & AMR ≥ 5% & EAS/SAS/AFR/OCE < 5%, NAT: subjects with EUR < 90% & AMR ≥ 60% & EAS < 20% & SAS < 15% & AFR/OCE < 5%, MIX: Uncategorized subjects. The LAT and NAT groups were combined into a single group with the overall LAT label.

### Statistics for CNV genome wide association

The association of deletions or duplications with case status was tested by logistic regression, controlling for confounding variables such as sex, genotyping platform, and 10 ancestry principal components (PCs) derived from SNP genotypes (**Model 1**). A logistic regression using only the covariates was used as a null model (**Null Model 1**). Datasets containing families used a conditional logistic regression with an extra covariate corresponding to the family ID (**Model 2, Null Model 2**). A chi-square test was then performed on the 2 models to obtain a p-value. A meta-analysis weighted by sample size (N-weighted) was applied across all platforms using METAL ^30^. Effective sample size (**N**_**eff**_) was used to determine the contribution of each platform. Associations were conducted at the breakpoint level since our sample size provided enough CNVs to obtain sufficient power. A breakpoint was included in the analysis if it contained at least 12 CNVs.

*Model* 1: *aff* ∼ *CNV* + *sex* + *PCs*

*Null Model* 1: *aff* ∼ *sex* + *PCs*

*Model* 2: *aff* ∼ *CNV* + *sex* + *PCs* + *strata*(*FID*)

*Null Model* 2: *aff* ∼ *sex* + *PCs* + *strata*(*FID*)

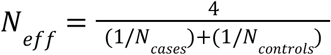

### Addressing statistical confounders in large-scale genome-wide meta-analysis of rare CNVs

In our experience, the key statistical confounders that must be addressed when performing a genome-wide meta-analysis of rare CNVs across many cohorts are (1) heterogeneity in CNV detection due to SNP genotyping platform and (2) The sparse data (zero cell) problem ^91^, where some loci produce summary statistics with very high standard errors in a subset of the cohort due to zero counts in cases or controls.

#### Heterogeneity of SNP genotyping platforms

Population stratification, a common confounder in genome-wide association studies ^92^ can be addressed in studies of rare variants by controlling for ancestry principal components derived from SNP genotypes ^93^. In large-scale collaborative studies of rare variants, however, there is another major confounder that must be addressed: differences in variant frequency between cohorts that is attributable to differences in the technology platforms used for rare variant detection.

“Platform stratification” is a confound that is conceptually similar to population stratification but is tied to genotyping/sequencing platform instead of ancestry. In large-scale collaborative studies of CNVs, where datasets from multiple cohorts are combined, differences in CNV frequencies between datasets can arise due to differences in CNV detection by the genotyping platforms that are used with each cohort. For instance, due to regional differences in probe coverage, a given CNV may be detected with greater sensitivity by platform A than by platform B. When two datasets genotyped with platforms A and B are combined, a false-positive association with the CNV can occur, particularly when the relative proportions of cases and controls differ between datasets (**Fig S3**). Thus “platform stratification” can artificially produce differences in CNV frequency between cases and controls.

To identify signals that are potentially attributable to platform stratification, we derived a measure for the platform or dataset specificity of CNV counts for a given probe. CNV frequency is calculated within-platform and within-dataset (**Equation 1**). A weighted deviance score (WDS) is then calculated for each platform/dataset (**Equation 2**) and a specificity index (SI) is derived by taking the maximum of the WDS across platforms/datasets (**Equation 3**). When calculating the SI score, all counts were used to calculate E_i_ in Equation 2, but only platforms/datasets with >2 CNVs were included when calculating the maximum in Equation 3. This prevented inaccurate SI scores being driven by single counts in smaller sample sizes. A threshold value of SI>0.2 was chosen for platform and SI>0.6 was chosen for dataset to flag probes that display platform stratification.

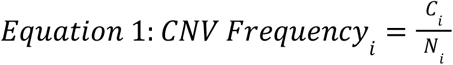

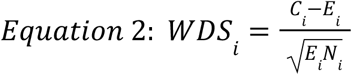

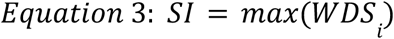

*E*_*i*_ = *pN*_*i*_ ; *N*_*i*_ : # *of samples on platform i*; 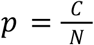

*C*_*i*_: # *of CNV counts on platform i*; *E*_*i*_: *Expected CNV counts on platform i*

*N*: # *of total samples*; *C*: # *of total CNV counts*

When association tests are performed across the combined SCZ sample in a single logistic regression model, the associations that arise cluster into groups based on the SI measure. Most associations in SCZ, including all well established “known” associations, are enriched among probes that have low SI. A second small cluster of probes can be seen with high SI (**Fig. S3A**). In a manhattan plot, these appear as distinct association peaks that persist even when platform is included as a covariate in the logistic regression model (**Fig. S3C**). These spurious signals are addressed by a meta-analysis of CNV summary statistics by platform as described below.

#### Sparse data problem

A major confounder in genome-wide association studies of rare variants is attributable to sparse counts of rare variants within subsets of the combined sample, where there can be zero values in cases and controls distributed across the cohort ^91^. These zero values can lead to greatly inflated standard error estimates for specific loci in a subset of the summary statistics used in the meta-analysis. When meta-analysis was weighted by standard error, tests that yielded high standard error estimates (**Fig S3B**, SEM>1) contributed to many spurious associations throughout the genome (**Fig. S3D**).

#### Sample size-weighted meta-analysis of samples grouped by genotyping platform

Platform heterogeneity was adequately addressed when (1) samples were first grouped by genotyping platform, and meta-analysis was performed across platforms and (2) meta-analysis was weighted by the samples sizes of each platform instead of standard error. Following this workflow, the combined association data does not show strong signals that are driven by probes subject to platform heterogeneity or by probes that show high SE estimates in the combined sample (**Fig. S3E**).

### Estimation of multiple test correction

The p-value threshold that gives a family-wise error rate (FWER) of 0.05 was calculated as an adjusted Bonferroni correction. The total number of tests was replaced with the total number of independent tests (Equation 4). Independent tests were counted after removing tests with >4% Jaccard index. The Jaccard index threshold was chosen by comparing multiple test corrections based on permutations in the SCZ cohort on chr1 with Bonferroni corrections at different Jaccard index thresholds (**Fig. S4**).

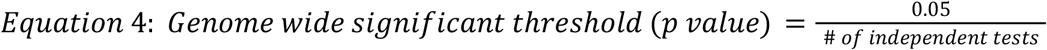

### Investigating the Pleiotropic effects of CNVs across 6 diagnostic categories

To investigate the full range of psychiatric conditions associated with each locus, we performed a more detailed characterization of the effect sizes of specific CNV alleles across the 6 diagnostic categories. For each locus with recurrent (NAHR) breakpoints (1q21.1-21.2, 3q29, 7q11.23, 15q11.2, 15q11.2-13.1, 15q13.1, 15q13.3, 16p13.11, 16p12.2, 16p11.2, 22q11.21), we identified all of the distinct alleles within each locus and genotyped each individually. For loci with non-recurrent (randomly distributed) breakpoints (ASTN2, NRXN1, SMYD3, USP7-HAPSTR1, 8p23, 22q13.3) we tested individual genes. Altogether, across the 18 regions listed above, there were 43 distinct loci, and DEL and DUP were tested for each (86 alleles, **Table S9**). Only tests with a count of at least 12 in the combined cases and controls were considered, thus ∼50 tests are reported for each diagnostic category.

### Annotating which CNV loci are recurrent (NAHR) or non-recurrent, and annotating clinically reportable CNVs

All of the associations reported in this study were “CNV hotspots”, i.e. loci that are prone to genomic instability. A majority were “NAHR” loci where recurrent de novo CNVs occur by non-allelic homologous recombination (NAHR) and de novo mutation events produce deletions and duplications with similar breakpoints ^57,94^. The remaining loci consisted of “fragile sites” where frequent double strand breaks give rise to many non-recurrent deletions and duplications that have breakpoints that are more randomly distributed across the locus. **Table S7** provides a guide to which CNV loci are NAHR or are non-recurrent. Where applicable, we also include links to clinical guidelines for CNVs that are recognized as pathogenic and reported in clinical genetic testing.

**Table S7** was prepared as follows. An initial “morbidity map” of the genome was created by Cooper et al. ^43^, identifying NAHR-mediated regions and characterizing their associations using large clinical microarray datasets from pediatric developmental disorder cases. This study has since become a standard reference list of known recurrent CNVs used by our group and others ^21^. We created an updated version of the morbidity map from the union of loci from Cooper et al.^43^, Kendall et al^21^, and this study. Breakpoints were refined to facilitate genotyping of distinct CNV alleles. A majority of the loci in the morbidity map are known CNVs that are routinely reported in clinical genetic testing. Where appropriate, we provide a URL link to clinical guidelines from Gene Reviews^95^, Clin Gen^96^, OMIM ^97^ or other databases. In addition, we label the CNV alleles corresponding to “GWS loci” and “Other Clinically-Reportable CNVs” in **Figure 2A**.

### Calling genotypes for each locus

NAHR CNVs were genotyped based on CNV calls with >50% reciprocal overlap with the locus. For complex loci with several breakpoints and multiple subregions (BP1-BP2, BP2-BP3, BP3-BP4…), such as 15q11-13 and 22q11, each subregion was genotyped using one-way overlap with CNVs, and specific alleles were called based on the subregions that were spanned by each CNV. For example, a DEL that spans 22q11.21 A-B, B-C, and C-D would be called “22q11.2 A-D”.

For single genes, we tested counts for predicted loss of function (LoF) variants (DELs that intersect with exons) and for predicted gain of function (DUPs that span at least one full-length isoform. For single gene loci we also compared associations for intronic DELs and DELs predicted to cause LoF or an in-frame deletion (**Fig. 4**).

### Testing association of genome-wide CNV burden

The CNV burden of deletions and duplications was tested by logistic regression, controlling for confounding variables such as sex, genotyping platform, and 10 ancestry principal components (PCs) derived from SNP genotypes. CNV burden was calculated for each platform to control for differences in probe coverage between different cohorts. SE-weighted meta-analysis was used to estimate CNV burden using METAL ^30^ since N-weighted meta-analysis does not provide effect sizes.

For analysis of genome-wide CNV burden, the number of base pairs that overlapped a CNV genome-wide was used as the independent variable (**Burden Model 1**) and CNV burden was calculated by comparing it to a model that only contained covariates (**Null Model**). CNV burden was also calculated for CNVs > 1Mb in size with the same approach (**Burden Model 2, Null Model**). Genes were partitioned into two tiers according to their pLI score: high pLI (T1; pLI>0.5) and low pLI (T2; pLI ≤0.5). CNV burden was calculated for all genes (**Burden Model 3A, Gene Null Model**), Tier 1 genes (**Burden Model 3B, Gene Null Model**), and Tier 2 genes (**Burden Model 3C, Gene Null Model**) while controlling for the out-of-category genome-wide burden defined as the number of base pairs that do not overlap with the gene category being tested. CNV burden was calculated for the GWS loci (**Burden Model 4A, Null Model**), clinically-reportable CNVs (**Burden Model 4B, Null Model**), and GWS loci + clinically-reportable CNVs (**Burden Model 4C, Null Model**) categories by counting the number of CNVs an individual carried in each category.

*Burden Model* 1: *aff* ∼ # *of base pairs overlapping CNV* + *sex* + *PCs*

*Burden Model* 2: *aff* ∼ # *of base pairs overlapping CNV* > 1*Mb* + *sex* + *PCs*

*Burden Model* 3*A*: *aff* ∼ # *of base pairs overlapping genes and CNV* + *OOC* + *sex* + *PCs*

*Burden Model* 3*B*: *aff* ∼ # *of base pairs overlapping T*1 *genes and CNV* + *OOC* + *sex* + *PCs*

*Burden Model* 3*C*: *aff* ∼ # *of base pairs overlapping T*2 *genes and CNV* + *OOC* + *sex* + *PCs*

*Burden Model* 4*A*: *aff* ∼ # *of CNVs overlapping GWS loci* + *sex* + *PCs*

*Burden Model* 4*B*: *aff* ∼ # *of CNVs overlapping clinically reportable CNVs* + *sex* + *PCs*

*Burden Model* 4*C*: *aff* ∼ # *of CNVs overlapping GWS & clinically reportable CNVs* + *sex* + *PCs*

*Null Model*: *aff* ∼ *sex* + *PCs*

*Gene Null Model*: *aff* ∼ *OOC* + *sex* + *PCs*

### Estimation of CNV frequencies in each diagnostic category

As mentioned above, it is necessary to control for genotyping platform when estimating the relative frequencies of CNVs in cases and controls. Thus, the CNV frequencies in cases for each category in **Fig. 2A** were derived from the odds ratio estimates from meta-analysis of CNV counts. We use the observed control frequency and the odds ratio from meta-analysis of CNV burden across platforms (**Burden Model 4, Null Model, Table S6**) to produce an accurate estimate of CNV case frequency (**Equation 5**).

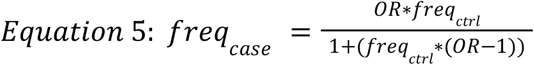

*freq*_*ctrl*_ : CNV frequency in the combined control sample

### Total variance explained by CNVs

The proportion of variance explained by CNVs was determined by calculating Nagelkerke’s R^2^ between a CNV model and a null model. Nagelkerke’s R^2^ was calculated for each platform separately to control for differences in probe coverage between different cohorts. Bootstrapping was implemented to estimate standard errors for the R^2^ estimate on each platform and summary statistics were combined through meta-analysis using the *metafor* package in R ^98^.

Nagelkerke’s R^2^ was calculated for genome-wide burden of length, loci that were genome-wide significant in our study, clinically-reportable CNVs, and the combination of all categories. The R^2^ estimate for genome-wide burden of length was tested by adding the length of all DELs and DUPs separately for each individual and comparing a logistic regression model of aggregate length and covariates (**Burden Model**) against a model with only covariates (**Null Model**). To estimate R^2^ for the GWS loci and clinically-reportable CNVs, a genotype matrix was created for each category (separate genotypes for DEL and DUP) and a logistic regression model with each locus genotype as a separate variable plus covariates (**Genotype Model**) was compared against a model with only covariates (**Null Model**). The genotype matrix for GWS loci and clinically-reportable CNVs was then combined and supplemented with the genome-wide burden annotations (DEL and DUP) to estimate the R^2^ for the combined genotype and genome-wide burden using the same method (**Combined model, Null Model**).

*Burden Model*: *aff* ∼ # *of base pairs overlapping DEL* + # *of base pairs overlapping DUP*

+ *sex* + *PCs*

*Genotype Model*: *aff* ∼ *CNVgenotype* + *sex* + *PCs*

*Combined Model*: *aff* ∼ *CNVgenotype* + # *of base pairs overlapping DEL*

+ # *of base pairs overlapping DUP* + *sex* + *PCs*

*Null Model*: *aff* ∼ *sex* + *PCs*

### Estimating effect sizes for CNVs at 18 loci across 6 diagnostic categories

Effect size estimates from N-weighted meta-analysis are scaled as a Z-score by default. To derive estimates of the odds ratio for each CNV, a mega-analysis was performed on the combined platforms, and odds ratio estimates were added to **Table S9**. For associations with high standard error (SEM>20) in the mega-analysis (due to zero counts in cases or controls), we applied a continuity correction to the effect size estimate similar to the Haldane-Anscombe correction for estimating odds ratios when there is a zero count in a contingency table ^99^ ^100^. To calculate the odds ratio of a CNV that contains a zero count in cases or controls, we added a count of 1 to each cell in the contingency table by randomly sampling 4 subjects from the cohort with replacement (duplicating 2 cases and 2 controls), then the appropriate genotypes were assigned to each (1 case with CNV, 1 case no CNV, 1 control with CNV, 1 control no CNV), and the four subjects were added to the sample.

### Assembling HiFi reads from samples carrying the SMYD3 duplication

HiFi long-read whole genome sequencing was performed on 3 samples using the PacBio Revio platform. Minimap2 v2.24 was used for alignment, DeepVariant v1.5 for variant calling, WhatsHap v2.0 for phasing and haplotagging reads, and Flye v2.9.2 for assembly.

## Supplemental Material

## Supplementary Figures

**Figure S1:**
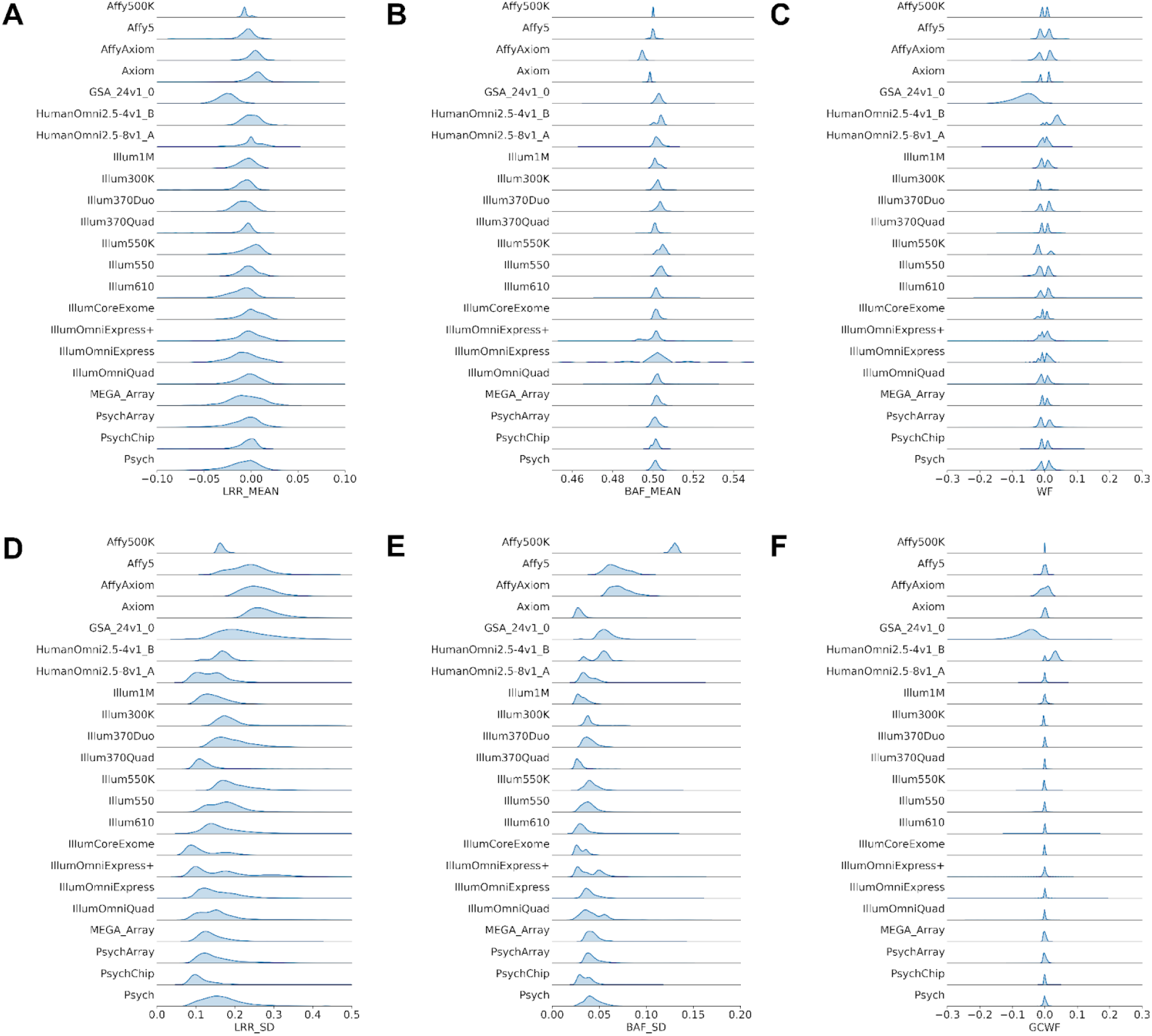
QC metrics across different platforms in the current PGC CNV study. For each platform, we show the distribution of each QC metric at the sample level including A) The mean Log R Ratio (LRR_MEAN), B) The mean B Allele Frequency (BAF_MEAN), C) Waviness Factor (WF) D) Log R Ratio standard deviation (LRR_SD), E) B Allele Frequency standard deviation (BAF_SD), and F) GC waviness factor (GCWF). LRR_SD measures the variability of LRR across a sample while BAF_SD measures the variability of BAF across a sample. Smaller standard deviation values lead to more accurate CNV calling. WF measures the total amount of fluctuation in signal intensity in a sample while GCWF measures the amount of signal intensity fluctuation explained by local GC content.

**Figure S2:**
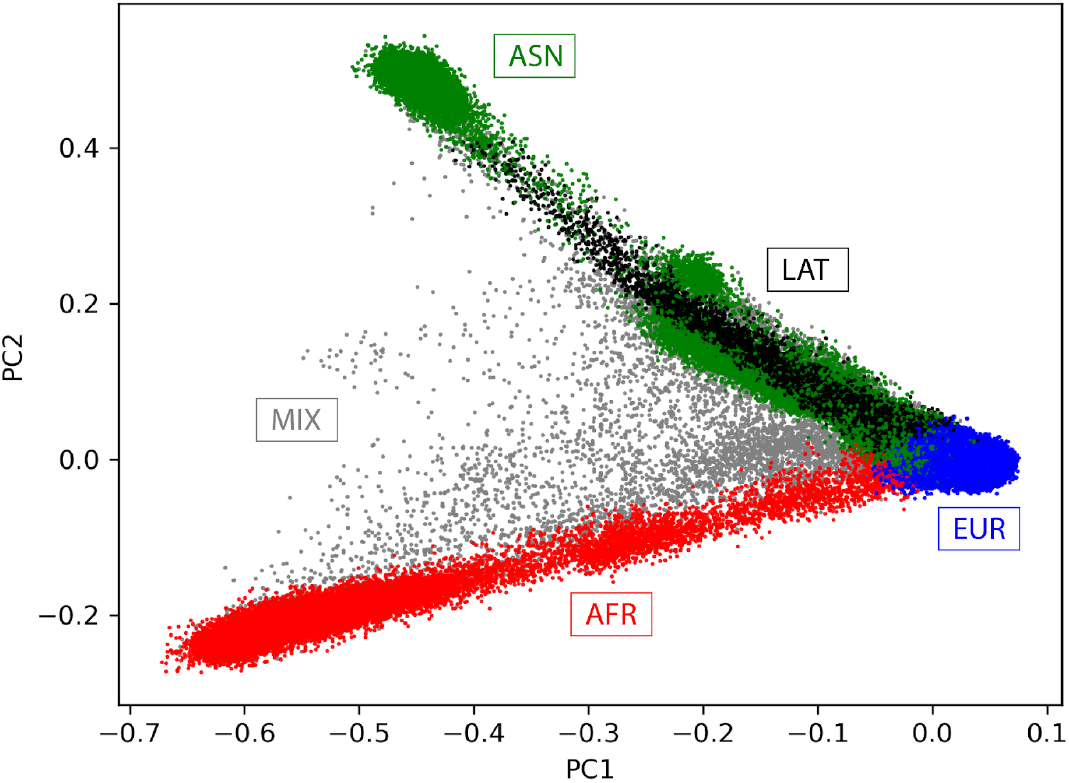
Sample groupings for all individuals in the current PGC CNV study.

**Figure S3:**
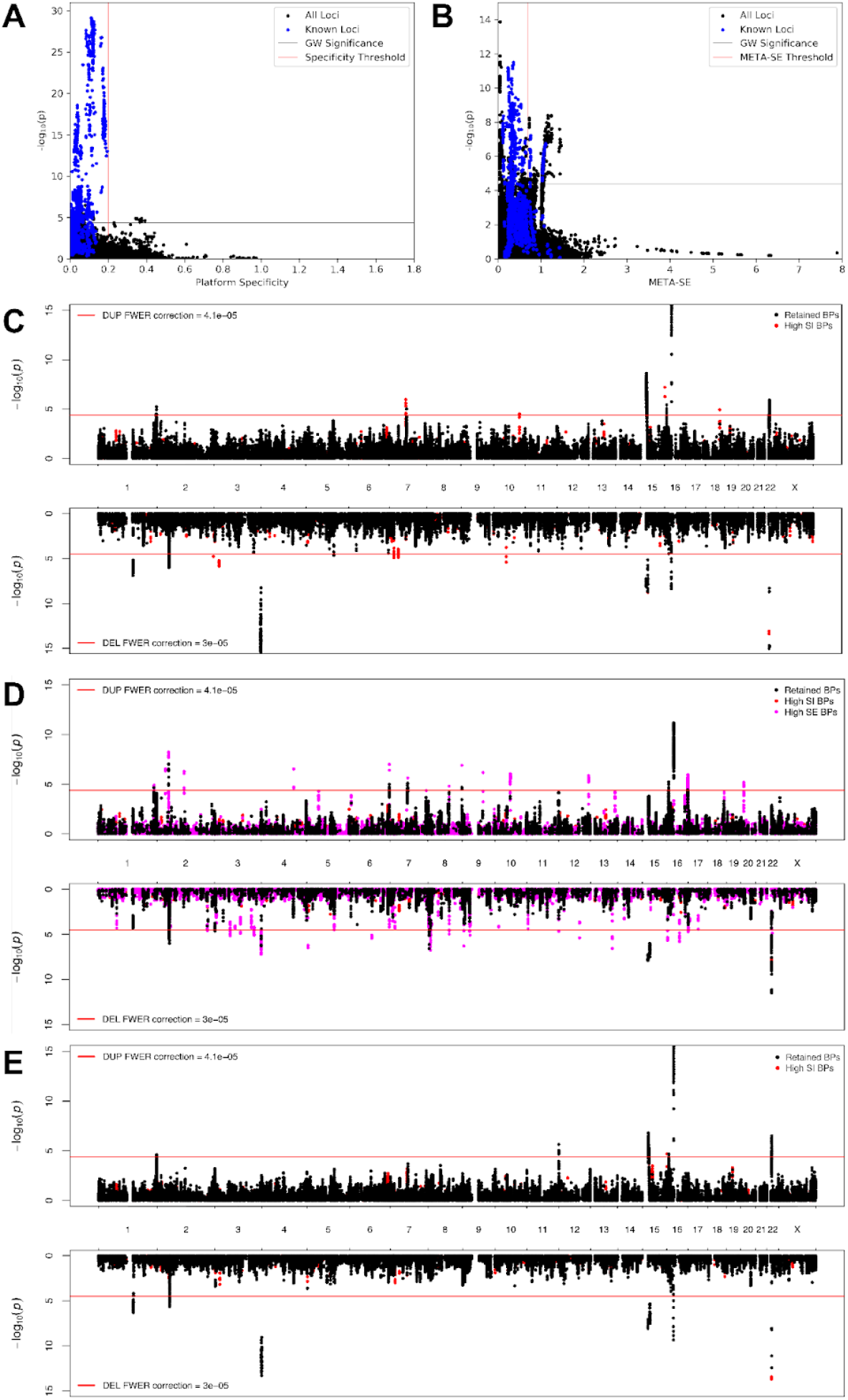
Platform/dataset specificity index filter and CNV GWAS for SCZ shows platform-specific spurious associations. -log_10_(pvalue) from the SCZ CNV GWAS vs. A) specificity index for platform and B) META-SE standard error are shown. Deletions and duplications are both included. Previously identified SCZ loci are shown in blue and the rest of the breakpoints are shown in black. The known loci cluster near lower specificity values and low SE while the spurious associations that need to be filtered cluster at high SI values and high SE. A threshold of 0.6 SI was chosen to filter out CNVs coming from a single platform and 0.2 SI for dataset. N-weighted meta-analysis was chosen instead of a threshold for META-SE. C) Mega-analysis CNV GWAS in SCZ controlling for genotyping platform as a covariate does not properly correct for genotyping platform. D) CNV GWAS using SE-weighted meta-analysis across platforms does not properly account for case/control imbalances in different platforms and produces spurious associations with high standard error. E) CNV GWAS using N-weighted meta-analysis correctly accounts for differences in genotyping platform and case/control imbalances. Breakpoints filtered by the platform/dataset specificity filter are shown in red. Breakpoints with high standard error are shown in magenta. Black are all retained breakpoints.

**Figure S4:**
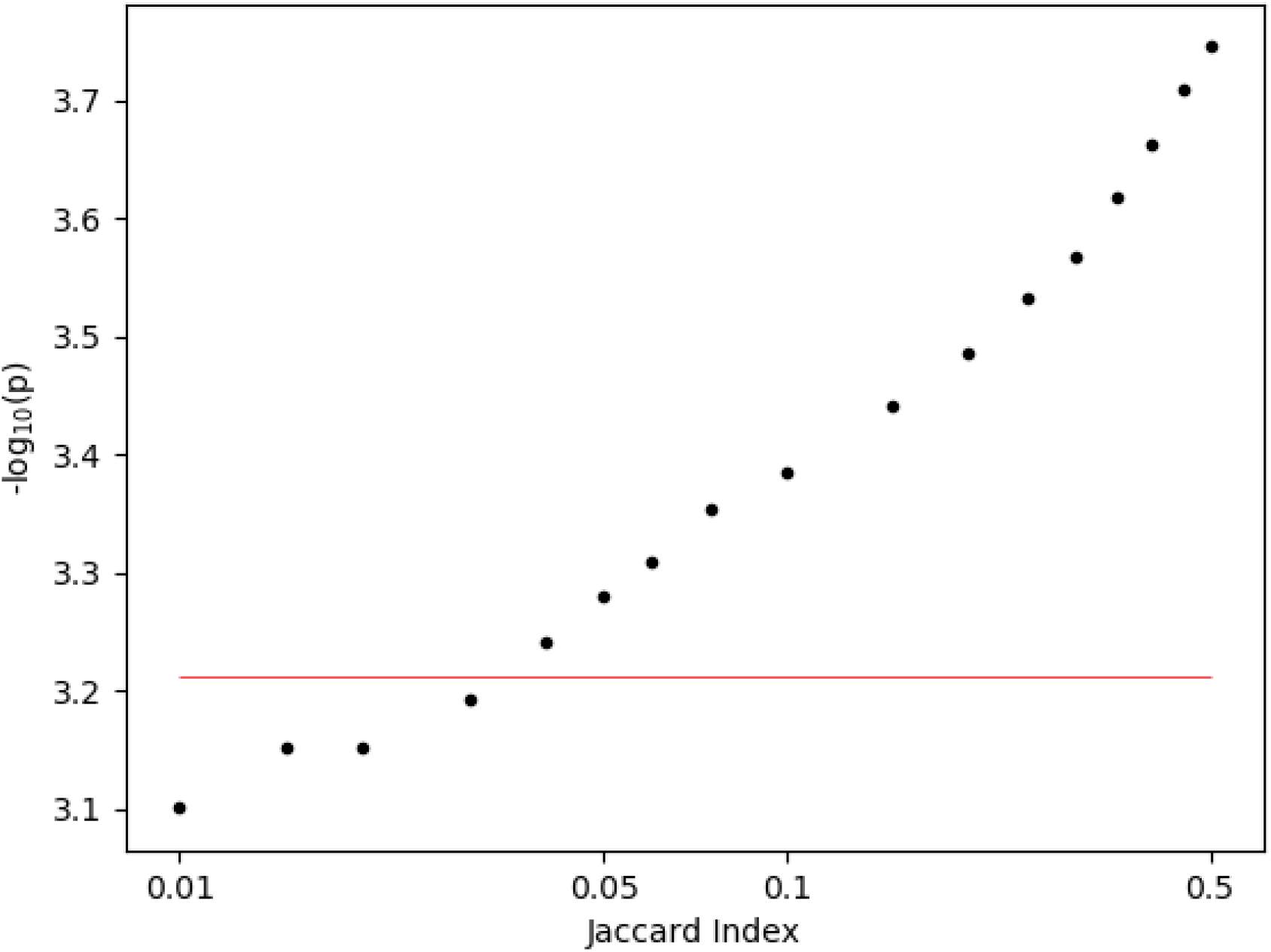
P-value thresholds for different values of Jaccard Index. The red line shows the p-value (6.14×10^−4^) that gives an FWER of 0.05 based on permutations in the SCZ cohort on chr1.

**Figure S5:**
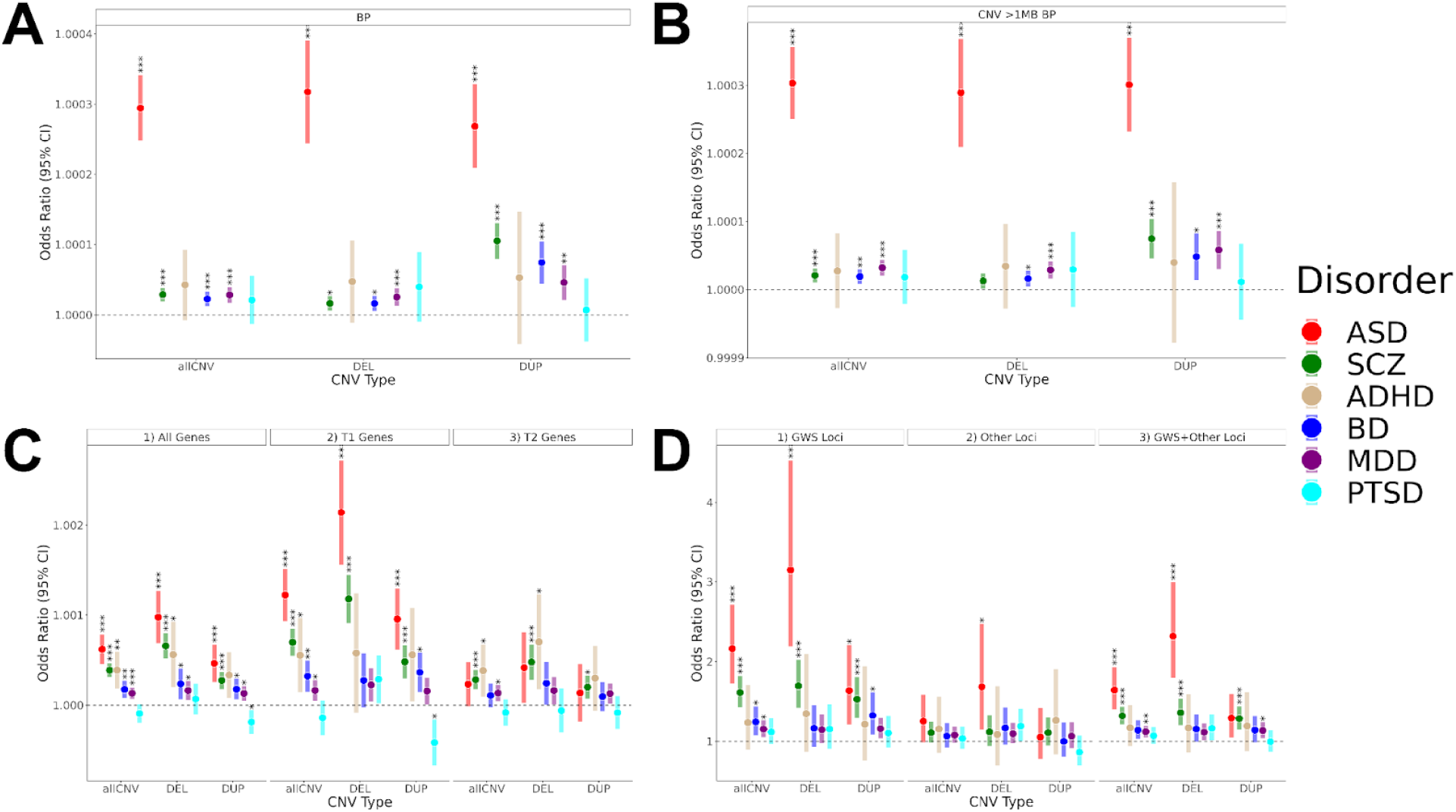
Effect sizes of rare CNVs differ by diagnosis, genes, and regions. A) A CNV burden test for all base pairs was performed for each diagnosis. Rare CNVs contribute to risk in most psychiatric conditions at different magnitudes. B) A CNV burden test was performed for large CNVs with length >1Mb. Results are very similar to all CNVs (Panel A). C) Genes were stratified by their probability of loss of function intolerance (pLI). Tier 1 Genes are defined as pLI > 0.5 and Tier 2 genes have pLI ≤ 0.5. CNVs overlapping Tier 1 genes are enriched in cases with the largest effects in ASD and SCZ. D) CNV burden was tested within the genome-wide significant (GWS) loci identified in this study, clinically-reportable CNVs that were not genome-wide significant in this study (Other loci), and in a combined group (GWS+Other loci) **Table S6**.

**Figure S6:**
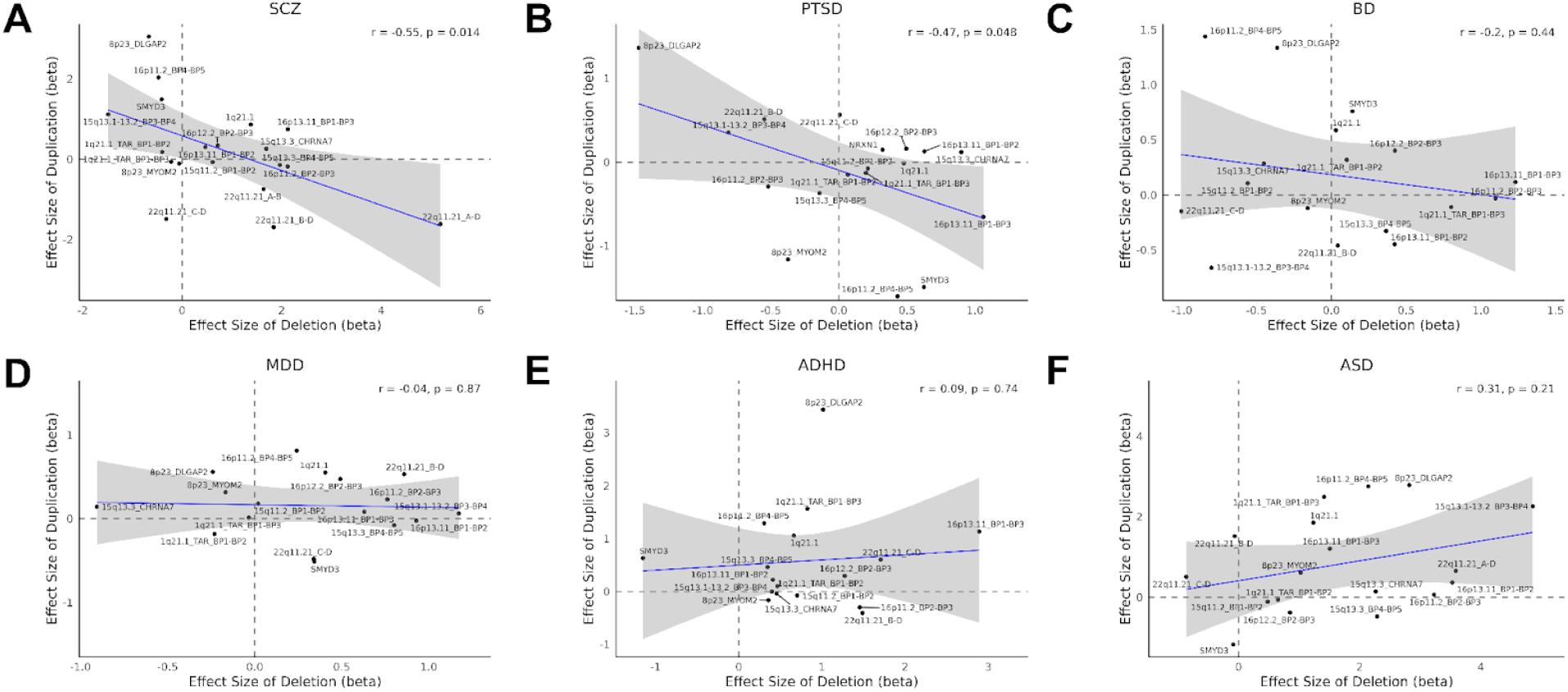
Dose-dependent effects of rare CNVs in different psychiatric traits. Effect sizes for duplications and deletions show a significant inverse dosage relationship for A) SCZ and B) PTSD. Dosage relationship curves are also shown for C) BD D) MDD E) ADHD and F) ASD. **Table S9**.

**Figure S7:**
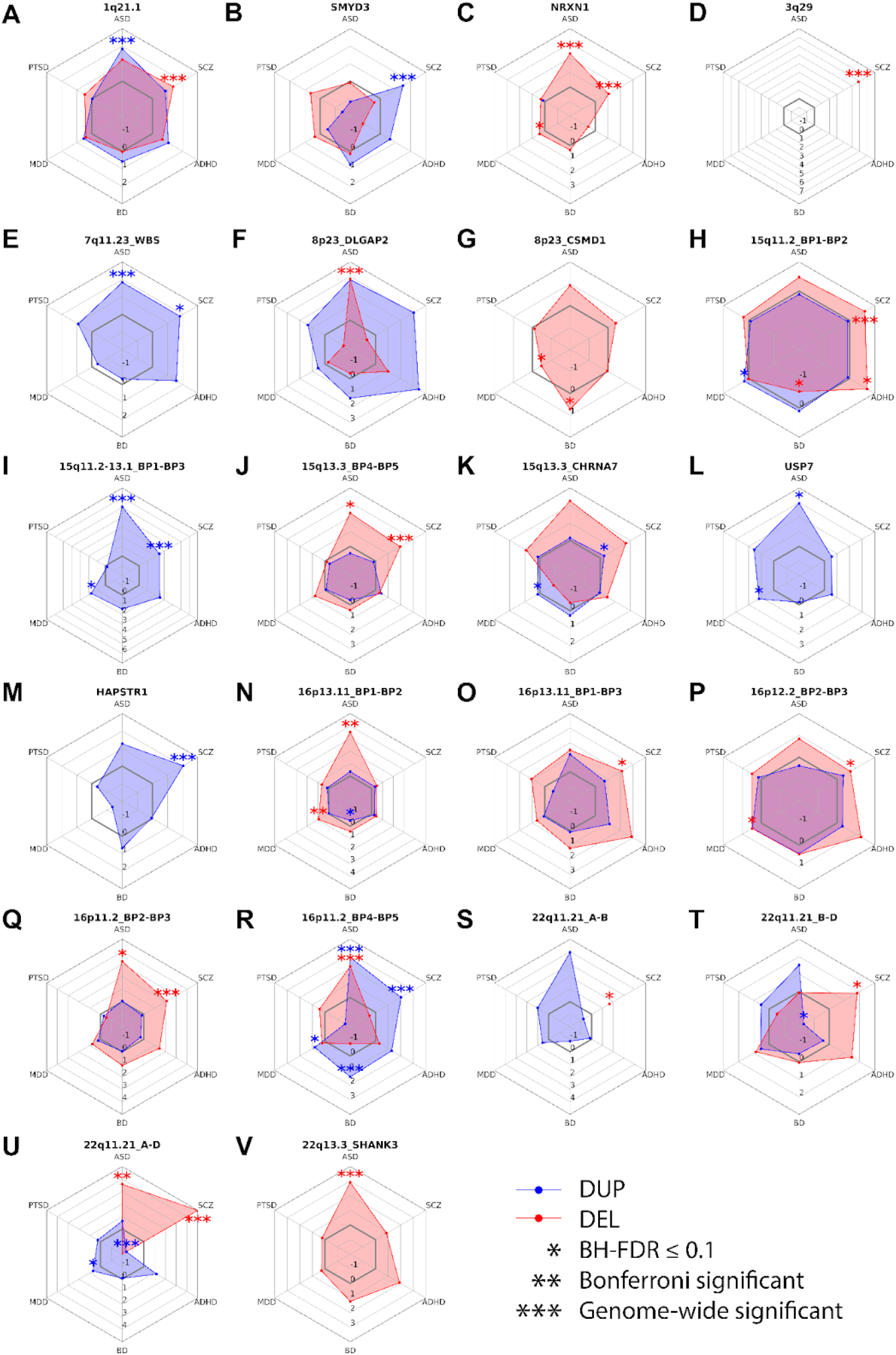
Effect Size across 6 diagnoses for each locus. Effect sizes were estimated by mega-analysis. For summary statistics with high SE, effect sizes were re-estimated with continuity correction. **Table S9**.

## References

1. Cross-Disorder Group of the Psychiatric Genomics Consortium et al. Genetic relationship between five psychiatric disorders estimated from genome-wide SNPs. Nat. Genet. 45, 984–994 (2013).

2. Schizophrenia Working Group of the Psychiatric Genomics Consortium. Biological insights from 108 schizophrenia-associated genetic loci. Nature 511, 421–427 (2014).

3. Malhotra, D. & Sebat, J. CNVs: harbingers of a rare variant revolution in psychiatric genetics. Cell 148, 1223–1241 (2012).

4. Weiner, D. J. et al. Polygenic architecture of rare coding variation across 394,783 exomes. Nature 614, 492–499 (2023).

5. Singh, T. et al. Rare coding variants in ten genes confer substantial risk for schizophrenia. Nature 604, 509–516 (2022).

6. Sebat, J. et al. Strong association of de novo copy number mutations with autism. Science 316, 445–449 (2007).

7. Walsh, T. et al. Rare structural variants disrupt multiple genes in neurodevelopmental pathways in schizophrenia. Science 320, 539–543 (2008).

8. International Schizophrenia Consortium. Rare chromosomal deletions and duplications increase risk of schizophrenia. Nature 455, 237–241 (2008).

9. Kendall, K. M. et al. Association of Rare Copy Number Variants With Risk of Depression. JAMA Psychiatry 76, 818–825 (2019).

10. Maihofer, A. X. et al. Rare copy number variation in posttraumatic stress disorder. Mol. Psychiatry 27, 5062–5069 (2022).

11. Pinto, D. et al. Convergence of genes and cellular pathways dysregulated in autism spectrum disorders. Am. J. Hum. Genet. 94, 677–694 (2014).

12. Kirov, G. et al. De novo CNV analysis implicates specific abnormalities of postsynaptic signalling complexes in the pathogenesis of schizophrenia. Mol. Psychiatry 17, 142–153 (2012).

13. Marshall, C. R. et al. Contribution of copy number variants to schizophrenia from a genome-wide study of 41,321 subjects. Nat. Genet. 49, 27–35 (2017).

14. Weiss, L. A. et al. Association between microdeletion and microduplication at 16p11.2 and autism. N. Engl. J. Med. 358, 667–675 (2008).

15. Sanders, S. J. et al. Insights into Autism Spectrum Disorder Genomic Architecture and Biology from 71 Risk Loci. Neuron 87, 1215–1233 (2015).

16. McCarthy, S. E. et al. Microduplications of 16p11.2 are associated with schizophrenia. Nat. Genet. 41, 1223–1227 (2009).

17. Kirov, G. et al. Neurexin 1 (NRXN1) deletions in schizophrenia. Schizophr. Bull. 35, 851–854 (2009).

18. Karayiorgou, M., Simon, T. J. & Gogos, J. A. 22q11.2 microdeletions: linking DNA structural variation to brain dysfunction and schizophrenia. Nat. Rev. Neurosci. 11, 402–416 (2010).

19. Stefansson, H. et al. Large recurrent microdeletions associated with schizophrenia. Nature 455, 232–236 (2008).

20. Sebat, J., Levy, D. L. & McCarthy, S. E. Rare structural variants in schizophrenia: one disorder, multiple mutations; one mutation, multiple disorders. Trends Genet. 25, 528–535 (2009).

21. Kendall, K. M. et al. Cognitive Performance Among Carriers of Pathogenic Copy Number Variants: Analysis of 152,000 UK Biobank Subjects. Biol. Psychiatry 82, 103–110 (2017).

22. Stefansson, H. et al. CNVs conferring risk of autism or schizophrenia affect cognition in controls. Nature 505, 361–366 (2014).

23. Qiu, Y. et al. Oligogenic Effects of 16p11.2 Copy-Number Variation on Craniofacial Development. Cell Rep. 28, 3320–3328.e4 (2019).

24. Owen, D. et al. Effects of pathogenic CNVs on physical traits in participants of the UK Biobank. bioRxiv (2018) doi:10.1101/355297.

25. Crawford, K. et al. Medical consequences of pathogenic CNVs in adults: analysis of the UK Biobank. J. Med. Genet. 56, 131–138 (2019).

26. Antaki, D. et al. A phenotypic spectrum of autism is attributable to the combined effects of rare variants, polygenic risk and sex. Nat. Genet. 54, 1284–1292 (2022).

27. Davies, R. W. et al. Using common genetic variation to examine phenotypic expression and risk prediction in 22q11.2 deletion syndrome. Nat. Med. 26, 1912–1918 (2020).

28. Cross-Disorder Group of the Psychiatric Genomics Consortium. Electronic address: & Cross-Disorder Group of the Psychiatric Genomics Consortium. Genomic relationships, novel loci, and pleiotropic mechanisms across eight psychiatric disorders. Cell 179, 1469–1482.e11 (2019).

29. Wellcome Trust Case Control Consortium et al. Genome-wide association study of CNVs in 16,000 cases of eight common diseases and 3,000 shared controls. Nature 464, 713–720 (2010).

30. Willer, C. J., Li, Y. & Abecasis, G. R. METAL: fast and efficient meta-analysis of genomewide association scans. Bioinformatics 26, 2190–2191 (2010).

31. Riggs, E. R. et al. Copy number variant discrepancy resolution using the ClinGen dosage sensitivity map results in updated clinical interpretations in ClinVar. Hum. Mutat. 39, 1650–1659 (2018).

32. ClinGen. Dosage Sensitivity. https://search.clinicalgenome.org/kb/gene-dosage.

33. Guo, R. & Haldeman-Englert, C. R. 1q21.1 recurrent deletion. in GeneReviews(®) (University of Washington, Seattle, Seattle (WA), 1993).

34. Mulle, J. G. et al. 3q29 recurrent deletion. in GeneReviews(®) (University of Washington, Seattle, Seattle (WA), 1993).

35. Chung, W. K., Herrera, F. F. & Simon’s Searchlight Foundation. Health supervision for children and adolescents with 16p11.2 deletion syndrome. Cold Spring Harb. Mol. Case Stud. 9, (2023).

36. Óskarsdóttir, S. et al. Updated clinical practice recommendations for managing children with 22q11.2 deletion syndrome. Genet. Med. 25, 100338 (2023).

37. Lionel, A. C. et al. Disruption of the ASTN2/TRIM32 locus at 9q33.1 is a risk factor in males for autism spectrum disorders, ADHD and other neurodevelopmental phenotypes. Hum. Mol. Genet. 23, 2752–2768 (2014).

38. Vrijenhoek, T. et al. Recurrent CNVs disrupt three candidate genes in schizophrenia patients. Am. J. Hum. Genet. 83, 504–510 (2008).

39. Hogart, A., Wu, D., LaSalle, J. M. & Schanen, N. C. The comorbidity of autism with the genomic disorders of chromosome 15q11.2-q13. Neurobiol. Dis. 38, 181–191 (2010).

40. Grozeva, D. et al. Rare copy number variants: a point of rarity in genetic risk for bipolar disorder and schizophrenia. Arch. Gen. Psychiatry 67, 318–327 (2010).

41. Charney, A. W. et al. Contribution of Rare Copy Number Variants to Bipolar Disorder Risk Is Limited to Schizoaffective Cases. Biol. Psychiatry 86, 110–119 (2019).

42. Rucker, J. J. H. et al. Phenotypic Association Analyses With Copy Number Variation in Recurrent Depressive Disorder. Biol. Psychiatry 79, 329–336 (2016).

43. Cooper, G. M. et al. A copy number variation morbidity map of developmental delay. Nat. Genet. 43, 838–846 (2011).

44. Xu, G., Strathearn, L., Liu, B., Yang, B. & Bao, W. Twenty-year trends in diagnosed attention-deficit/hyperactivity disorder among US children and adolescents, 1997-2016. JAMA Netw. Open 1, e181471 (2018).

45. Maenner, M. J. et al. Prevalence and characteristics of Autism spectrum disorder among children aged 8 years - Autism and Developmental Disabilities Monitoring network, 11 sites, United States, 2020. MMWR Surveill. Summ. 72, 1–14 (2023).

46. Grove, J. et al. Identification of common genetic risk variants for autism spectrum disorder. Nat. Genet. 51, 431–444 (2019).

47. Trubetskoy, V. et al. Mapping genomic loci implicates genes and synaptic biology in schizophrenia. Nature 604, 502–508 (2022).

48. Mullins, N. et al. Genome-wide association study of more than 40,000 bipolar disorder cases provides new insights into the underlying biology. Nat. Genet. 53, 817–829 (2021).

49. Howard, D. M. et al. Genome-wide meta-analysis of depression identifies 102 independent variants and highlights the importance of the prefrontal brain regions. Nat. Neurosci. 22, 343–352 (2019).

50. Nievergelt, C. M. et al. International meta-analysis of PTSD genome-wide association studies identifies sex- and ancestry-specific genetic risk loci. Nat. Commun. 10, 4558 (2019).

51. Demontis, D. et al. Discovery of the first genome-wide significant risk loci for attention deficit/hyperactivity disorder. Nat. Genet. 51, 63–75 (2019).

52. Hamad, A. et al. Expanding the phenotypic spectrum of Chromosome 16p13.11 microduplication: A multicentric analysis of 206 patients. Eur. J. Med. Genet. 66, 104714 (2023).

53. Woodward, K. J. et al. Atypical nested 22q11.2 duplications between LCR22B and LCR22D are associated with neurodevelopmental phenotypes including autism spectrum disorder with incomplete penetrance. Mol. Genet. Genomic Med. 7, e00507 (2019).

54. Rees, E. et al. Evidence that duplications of 22q11.2 protect against schizophrenia. Mol. Psychiatry 19, 37–40 (2014).

55. Liu, W., Liu, F., Xu, X. & Bai, Y. Replicated association between the European GWAS locus rs10503253 at CSMD1 and schizophrenia in Asian population. Neurosci. Lett. 647, 122–128 (2017).

56. Engchuan, W. et al. Psychiatric disorders converge on common pathways but diverge in cellular context, spatial distribution, and directionality of genetic effects. medRxiv 2025.07.11.25331381 (2025).

57. Mefford, H. C. & Eichler, E. E. Duplication hotspots, rare genomic disorders, and common disease. Curr. Opin. Genet. Dev. 19, 196–204 (2009).

58. Lopes, I., Altab, G., Raina, P. & de Magalhães, J. P. Gene Size Matters: An Analysis of Gene Length in the Human Genome. Front. Genet. 12, 559998 (2021).

59. He, X. et al. Integrated model of de novo and inherited genetic variants yields greater power to identify risk genes. PLoS Genet. 9, e1003671 (2013).

60. Helmrich, A., Ballarino, M. & Tora, L. Collisions between replication and transcription complexes cause common fragile site instability at the longest human genes. Mol. Cell 44, 966–977 (2011).

61. Sarni, D. et al. 3D genome organization contributes to genome instability at fragile sites. Nat. Commun. 11, 3613 (2020).

62. Mortazavi, M. et al. Long-read genome sequencing in clinical psychiatry: RFX3 haploinsufficiency in a hospitalized adolescent with autism, intellectual disability, and behavioral decompensation. Am. J. Psychiatry appiajp20240471 (2025).

63. Modenato, C. et al. Effects of eight neuropsychiatric copy number variants on human brain structure. Transl. Psychiatry 11, 399 (2021).

64. Lin, A. et al. Reciprocal Copy Number Variations at 22q11.2 Produce Distinct and Convergent Neurobehavioral Impairments Relevant for Schizophrenia and Autism Spectrum Disorder. Biol. Psychiatry 88, 260–272 (2020).

65. Hippolyte, L. et al. The Number of Genomic Copies at the 16p11.2 Locus Modulates Language, Verbal Memory, and Inhibition. Biol. Psychiatry 80, 129–139 (2016).

66. Nguyen, T.-D. et al. Genetic heterogeneity and subtypes of major depression. Mol. Psychiatry 27, 1667–1675 (2022).

67. Jacquemont, S. et al. Genes To Mental Health (G2MH): A Framework to Map the Combined Effects of Rare and Common Variants on Dimensions of Cognition and Psychopathology. Am. J. Psychiatry 179, 189–203 (2022).

68. Zhang, Y., Li, C. & Yang, Z. Is MYND domain-mediated assembly of SMYD3 complexes involved in calcium dependent signaling? Front. Mol. Biosci. 6, 121 (2019).

69. Bernard, B. J., Nigam, N., Burkitt, K. & Saloura, V. SMYD3: a regulator of epigenetic and signaling pathways in cancer. Clin. Epigenetics 13, 45 (2021).

70. Williams, J. B. et al. Inhibition of histone methyltransferase Smyd3 rescues NMDAR and cognitive deficits in a tauopathy mouse model. Nat. Commun. 14, 91 (2023).

71. Fabini, E., Manoni, E., Ferroni, C., Rio, A. D. & Bartolini, M. Small-molecule inhibitors of lysine methyltransferases SMYD2 and SMYD3: current trends. Future Med. Chem. 11, 901–921 (2019).

72. Sanders, S. J. et al. Multiple recurrent DE Novo CNVs, including duplications of the 7q11.23 Williams syndrome region, are strongly associated with autism. Neuron 70, 863–885 (2011).

73. Uhlén, M. et al. Proteomics. Tissue-based map of the human proteome. Science 347, 1260419 (2015).

74. Qiao, H., Tian, Y., Huo, Y. & Man, H.-Y. Role of the DUB enzyme USP7 in dendritic arborization, neuronal migration, and autistic-like behaviors in mice. iScience 25, 104595 (2022).

75. Chen, H. et al. The Hao-Fountain syndrome protein USP7 regulates neuronal connectivity in the brain via a novel p53-independent ubiquitin signaling pathway. Cell Rep. 44, 115231 (2025).

76. Zhang, X.-W. et al. Neuroinflammation inhibition by small-molecule targeting USP7 noncatalytic domain for neurodegenerative disease therapy. Sci. Adv. 8, eabo0789 (2022).

77. Amici, D. R. et al. C16orf72/HAPSTR1 is a molecular rheostat in an integrated network of stress response pathways. Proc. Natl. Acad. Sci. U. S. A. 119, e2111262119 (2022).

78. Debs, S. R., Rothmond, D. A., Zhu, Y., Weickert, C. S. & Purves-Tyson, T. D. Molecular evidence of altered stress responsivity related to neuroinflammation in the schizophrenia midbrain. J. Psychiatr. Res. 177, 118–128 (2024).

79. Kushima, I. et al. Cross-disorder analysis of genic and regulatory copy number variations in bipolar disorder, schizophrenia, and autism spectrum disorder. Biol. Psychiatry 92, 362–374 (2022).

80. Wilson, P. M., Fryer, R. H., Fang, Y. & Hatten, M. E. Astn2, a novel member of the astrotactin gene family, regulates the trafficking of ASTN1 during glial-guided neuronal migration. J. Neurosci. 30, 8529–8540 (2010).

81. Behesti, H. et al. ASTN2 modulates synaptic strength by trafficking and degradation of surface proteins. Proc. Natl. Acad. Sci. U. S. A. 115, E9717–E9726 (2018).

82. The Schizophrenia Psychiatric Genome-Wide Association Study (GWAS) Consortium. Genome-wide association study identifies five new schizophrenia loci. Nat. Genet. 43, 969–976 (2011).

83. Coleman, J. R. I. et al. The genetics of the mood disorder spectrum: Genome-wide association analyses of more than 185,000 cases and 439,000 controls. Biol. Psychiatry 88, 169–184 (2020).

84. Stahl, E. A. et al. Genome-wide association study identifies 30 loci associated with bipolar disorder. Nat. Genet. 51, 793–803 (2019).

85. Network and Pathway Analysis Subgroup of Psychiatric Genomics Consortium. Psychiatric genome-wide association study analyses implicate neuronal, immune and histone pathways. Nat. Neurosci. 18, 199–209 (2015).

86. Manickam, K. et al. Exome and genome sequencing for pediatric patients with congenital anomalies or intellectual disability: an evidence-based clinical guideline of the American College of Medical Genetics and Genomics (ACMG). Genet. Med. 23, 2029–2037 (2021).

87. Schaefer, G. B., Mendelsohn, N. J. & Professional Practice and Guidelines Committee. Clinical genetics evaluation in identifying the etiology of autism spectrum disorders: 2013 guideline revisions. Genet. Med. 15, 399–407 (2013).

88. Auwerx, C. et al. Rare copy-number variants as modulators of common disease susceptibility. Genome Med. 16, 5 (2024).

89. Abraham, G., Qiu, Y. & Inouye, M. FlashPCA2: principal component analysis of Biobank-scale genotype datasets. Bioinformatics 33, 2776–2778 (2017).

90. Chen, C.-Y. et al. Improved ancestry inference using weights from external reference panels. Bioinformatics 29, 1399–1406 (2013).

91. Subbiah, M. & Srinivasan, M. R. Classification of 2×2 sparse data sets with zero cells. Stat. Probab. Lett. 78, 3212–3215 (2008).

92. Price, A. L., Zaitlen, N. A., Reich, D. & Patterson, N. New approaches to population stratification in genome-wide association studies. Nat. Rev. Genet. 11, 459–463 (2010).

93. Zhang, Y., Guan, W. & Pan, W. Adjustment for population stratification via principal components in association analysis of rare variants. Genet. Epidemiol. 37, 99–109 (2013).

94. Lupski, J. R. & Stankiewicz, P. Genomic disorders: molecular mechanisms for rearrangements and conveyed phenotypes. PLoS Genet. 1, e49 (2005).

95. Adam, M. P. et al. GeneReviews(®). (University of Washington, Seattle, Seattle (WA), 1993).

96. Clinical Genome Resource. Welcome to ClinGen. https://clinicalgenome.org/.

97. Home - OMIM. https://www.omim.org/.

98. Viechtbauer, W. Conducting Meta-Analyses in R with the metafor Package. J. Stat. Softw. 36, 1–48 (2010).

99. Haldane, J. B. S. The mean and variance of the moments of chi-squared when used as a test of homogeneity, when expectations are small. Biometrika 29, 133–134 (1940).

100. Anscombe, F. J. On estimating binomial response relations. Biometrika 43, 461–464 (1956).

